# Navigating the Impact of the 21st Century Cures Act Final Rule: A National Cross-Sectional Survey of U.S. Genetic Counselors

**DOI:** 10.1101/2024.10.13.24315353

**Authors:** Chenery Lowe, Laura Duncan, Victoria Morris, Katherine Anderson, Laynie Dratch, Debra J.H. Mathews

**Affiliations:** Stanford University, Center for Biomedical Ethics, Department of Pediatrics, Stanford, CA; Mayo Clinic, Center for Individualized Medicine, Jacksonville, FL.; Boston Children’s Hospital, Division of Genetics and Genomics, Boston, MA.; Vanderbilt University Medical Center, Division of Cardiovascular Medicine, Nashville, TN.; University of Pennsylvania, Penn Frontotemporal Degeneration Center and Department of Neurology, Philadelphia, PA.; Johns Hopkins University, Berman Institute of Bioethics, Department of Genetic Medicine, Baltimore, MD.

**Keywords:** 21st Century Cures Act Final Rule, Genetic Counseling, Clinical Documentation, Healthcare Policy, Genetic Results, Patient Harms

## Abstract

The 21^st^ Century Cures Act Final Rule (Final Rule) increases patient access to their health records but raises concerns about distress and misunderstanding of automatically released results and documentation. Little is known about genetic counselors’ (GCs’) experiences with the Final Rule. In Fall 2023, we conducted a cross-sectional survey of U.S. GCs about the Final Rule’s perceived impact on practice and assessed if being the ordering versus non-ordering provider for genetic testing affected perceptions of Final Rule-related changes. GCs (n=102) reported demographic and workplace characteristics, institutional policy changes, workflow changes, and perceived patient harms and benefits due to the Final Rule. To compare ordering and non-ordering providers, we conducted Fisher’s exact tests for categorical variables and trend tests for ordinal variables. Open response questions elicited examples of positive and negative patient impacts, the effects of patient characteristics, and practice changes in response to the Final Rule. Twenty-seven GCs were ordering providers and 66 were non-ordering providers. Relative to ordering providers, non-ordering providers expressed stronger agreement with statements indicating concern about the emotional impact on patients reviewing results or notes without support (P=0.002, 33% vs. 51% strongly agree), patients misunderstanding or misinterpreting results (P=0.025, 44% vs. 70% strongly agree), and patients contacting the inappropriate party to discuss results (P=0.023, 19% vs. 48% strongly agree). Agreement with the statement “patients have more knowledge/context/questions at our disclosure session due to previous results review” also differed by ordering provider status (P=0.007). In open responses, GCs expressed concerns about patients’ strong emotional reactions, patients misinterpreting results, workflow disruptions, and widening health disparities. Benefits included patients’ ability to be reassured, informed, or empowered earlier; ease of sharing health information; and more efficient workflow due to automatic release of results. These results emphasize the importance of clear communication within health systems and between patients and providers.

**What is known about this topic:** The 21^st^ Century Cures Act Final Rule (Final Rule) increases patients’ access to their health records, including automatically released results and clinical documentation. In prior studies, physicians and physician-led care team members recognize potential benefits for patient empowerment and potential harms due to information overload and misunderstanding, but little is known about mid-level providers’ perspectives, including genetic counselors.

**What this paper adds to the topic:** In a national survey, U.S. genetic counselors expressed worries about direct release of genetic test results, including concerns about patients’ emotional responses, risks related to misinterpretation of results and next steps, and the impact on their workflow. The study offers suggestions on how GCs can navigate changes imposed by the Final Rule.

## Introduction

The United States 21^st^ Century Cures Act (Cures Act), signed into law in 2016, builds on existing legislation to increase patient involvement in their medical care and research (“21st Century Cures Act, H.R. 34, 114th Congress,” 2015). As of April 2021, patients are entitled to immediate electronic access to their medical records, including clinical documentation notes from genetic counseling visits and genetic testing results, with limited exceptions.

Because the Final Rule allows patients to access their health information directly, medical providers may not have the chance to control how, when, or what information is communicated. It is therefore not surprising that early data regarding the impact of the Final Rule have recurrently commented on both patient perceptions and provider workflow (Brooks et al., 2023; Bruno et al., 2022; Congelosi et al., 2023; Mehan et al., 2021; Steitz et al., 2023; Sullivan et al., 2022). Studies across specialties, including neonatology, radiology, and oncology, have reported provider concerns about information overload for patients, including the access of provider-to-provider correspondence not intended for patient review, patient misunderstandings, and psychological impact of results on patients (Brooks et al., 2023; Mehan et al., 2021; Sullivan et al., 2024). A mixed methods study surveying neonatal providers in one NICU before and after the implementation of the Final Rule concluded that providers perceived that patients experienced benefits, such as increased parental involvement and engagement in their child’s care, though providers also had increasing concerns for information overload or misunderstanding results without clinician involvement, with specific concerns raised for brain imaging and genetic testing results (Sullivan et al., 2024).

In contrast to the concerns raised by medical providers, a study across several major United States (U.S.) medical centers found that most patients did not report an increase in worry with automatic release of either normal or abnormal results, including genetic testing results (Steitz et al., 2023). Moreover, patients and clinicians endorse personalization of results release based on patient preferences, testing type, and/or the results of testing as a means of reducing potential harm or distress to the patient (Brooks et al., 2023; Bruno et al., 2022).

To date, limited research has examined the perspectives of genetic counselors, who are trained specifically in effective communication of genetic testing results (Cragun & Zierhut, 2018; Zhong et al., 2020). Studies have demonstrated the benefits of the involvement of a genetic counselor in the results disclosure process, a process that is disrupted if patients access their results directly from an online portal (Patel et al., 2021). Of note, current clinical genetic reports are written beyond the college reading level (Davis et al., 2021), which may be inaccessible to patients of lower health literacy, particularly when high numeracy skills are needed to accurately assess risk. Moreover, while prior studies have primarily focused on physicians’ and physician-led care teams’ experiences and perspectives on the Final Rule, these findings may not be generalizable to genetic counselors, who are mid-level providers whose scope of practice includes independent interpretation and communication of genetic testing results (National Society of Genetic Counselors, n.d.). Additionally, in some states and institutions, genetic counselors may not be permitted to be the ordering provider for genetic testing, and therefore may not receive the test results directly, creating a delay between when the patient receives genetic results and when the genetic counselor is able to review and interpret the test report.

Here we use a multimethod survey to explore U.S. genetic counselors’ reactions to the implementation of the 21^st^ Century Cures Act Final Rule. Our primary aim was to examine how the Final Rule has affected U.S. genetic counselors’ practice, including how genetic counselors have adapted to the implementation of the Final Rule. Some genetic testing processes notify only the patient and ordering provider of completed results, which could introduce additional complexity for genetic counselors who coordinate testing and return results, but who are not considered the ordering provider of record. Thus, a secondary aim was to explore whether being the ordering provider for genetic testing versus being a non-ordering provider affected perceptions of the Final Rule’s impact on patients and patient care.

## Methods

### Participants

Genetic counselors who practiced in the U.S. after the implementation of the Final Rule were invited to participate in a survey regarding the Final Rule. An invitation to participate in the study was shared as part of a Final Rule plenary session at the National Society of Genetic Counselors (NSGC) Annual Conference and via NSGC research listserv. Participants were eligible for the study if they practiced as a genetic counselor in the U.S. after April 2021 and if they could read and answer survey questions in English. The survey was available from October 18, 2023 until December 31, 2023. Informed consent was obtained prior to start of the survey. This study was approved as exempt by the Mayo Clinic Institutional Review Board (23-007555).

### Instrumentation

This was multimethod survey developed by the authors as no validated measures exist for this topic. The survey items were primarily quantitative to facilitate assessment of the broad impact of the Final Rule across a range of workplace settings in the U.S. However, due to the limited prior research on the Final Rule’s impact on genetic counselors, we recognized that it would be valuable to include several open-ended questions that could help to contextualize quantitative findings and identify insights that our pre-specified survey questions would not have captured. The survey included defined quantitative response items and four open text boxes to elaborate on each topic in response to the probes described in more detail below. No further probing was conducted.

Participant demographics were collected including age, gender identity, race, ethnicity, years of experience, and participant work setting including NSGC geographic region of work, if their position was patient-facing, ordering provider status, employment setting, and specialty. Questions related to the Final Rule assessed institutional policies, resources, and training; and perceived impact of the Final Rule on workplace practices and on patients. The survey was administered via REDCap and the survey instrument is available as supplemental data.

For the items pertaining to institutional policies, resources, and training, respondents were presented with a list of possible policies, resources, or training, for example: “Due to the Final Rule, my workplace has institutional level policies for results/notes release.” Respondents could select between the following response categories: “This exists and a GC [genetic counselor] was involved,” “This exists and a GC was NOT involved,” “This exists and I don’t know if a GC was involved,” “This DOES NOT exist,” “I am not sure”, and “Not applicable at my workplace.”

For the items pertaining to the perceived impact of the Final Rule on workplace practices and on patients, respondents were presented with a statement related to possible perceived impacts of the Final Rule (e.g., “Due to the implementation of the Final Rule, I have less time to review results before discussing with a patient.”). Respondents were asked to select their level of agreement with the statement from the options “Strongly disagree,” “Disagree,” “Neither agree nor disagree,” “Agree,” “Strongly agree”, and “Not applicable for my position.”

Free text boxes allowed participants to provide specific examples in response to four open-response probes:

- Positive impact: “Please describe any examples you wish to share of how the Final Rule has positively impacted your patients/clients.”
- Negative impact: “Please describe any examples you wish to share of how the Final Rule has negatively impacted your patients/clients.”
- Patient characteristics: “Are there any demographics or characteristics of the population you typically work with that influence your perspective of the Final Rule (potential impact of patients having near-immediate access to their notes and results)? Please elaborate.”
- Practice changes: “If you practiced prior to the implementation of the Final Rule (April 2021), please describe any examples you wish to share about how the Final Rule has changed your personal practice as a genetic counselor including any changes you’ve made to consenting, documentation, patient results release, patient discussions, client conversations, report writing, etc.”

All four open-response probes were presented following the quantitative survey items and before the demographic questionnaire at the end of the survey.

### Procedures

Survey responses were reviewed for completion prior to analysis. Surveys that did not include responses to any demographic data or completion of the matrices assessing resources and genetic counselor involvement in implementation, workflow changes, and perceived patient impact were removed from analysis. Individuals who did not meet inclusion criteria and/or provide consent were prompted to end survey and therefore were not included in analysis.

### Data Analysis

#### Quantitative analyses

We conducted descriptive analyses to summarize the distribution of the variables of interest. As our objectives were primarily descriptive and there was no prior literature to inform effect sizes for the novel measures that we developed, we did not perform sample size calculations. For categorical outcomes, we reported the total frequency and percent of responses in each response category. To compare characteristics that were associated with being an ordering versus a non-ordering provider, we conducted Fisher’s exact test for non-ordered categorical variables due to low counts within some categories. For ordered categorical variables (age and years of experience), we conducted trend tests for ordinal variables. All analyses were conducted in R (Version 4.3) (R Core Team, 2023). Levels of missing data were low: other than the demographic questionnaire, none of the quantitative survey items had missing responses. Therefore, no form of imputation was used.

#### Qualitative analyses

Survey responses that included at least one response to an open-ended question were read by one coder (CL) and non-informative responses (e.g., “Na”, “-”) were excluded from analysis. Responses to each of the four open-ended questions were analyzed using inductive content analysis (Vears & Gillam, 2022). Multiple and overlapping codes were used when a participant provided more than one example or concept in their response. After completing open coding for all four questions, all authors reviewed the initial code system, code definitions, and examples to suggest minor revisions. Some suggested revisions included: expanding a code for patient emotions to include multiple subcodes and adding additional subcodes to classify different ways in which patients misunderstood their results.

Two coders (CL and KA) then applied the revised coding system across all four questions to capture examples that fit code definitions that were originally developed for responses to a different question – for instance, when respondents provided examples of negative impacts of the Final Rule in the response to the question about practice changes. Discrepancies regarding code assignment were resolved through discussion between the two coders and (if necessary) the full author group.

#### Integration of qualitative and quantitative analyses

The study followed a concurrent multimethod design in which the quantitative strand was given priority (“Distinguishing Mixed Methods Designs,” 2017, pp. 61–62). Due to the survey’s cross-sectional nature we were unable to probe participants’ responses, so a fixed design (Meissner et al., 2011) was used. The open-ended responses were initially open-coded without consideration of their relationship to the corresponding quantitative survey measures. In subsequent stages of analysis, we referred to the quantitative survey measures to determine ways in which the coded statements provided additional nuance to the quantitative survey items and ways in which they identified issues beyond the scope and assumptions of the quantitative items. These insights guided our approach to sub-category formation and identification of categories that highlighted implementation issues or solutions that were not included in the quantitative measures.

## Results

### Quantitative survey results

#### Participant characteristics

Of the 234 surveys started, 102 participants were included in analysis; 31 did not meet inclusion criteria and/or provide consent and 101 did not complete matrices and demographic data. Ninety-two participants (90%) identified as women, 7 (7%) as men, 1 (1%) did not provide a gender, and 2 (2%) preferred not to say their gender. Eighty-seven participants reported their race as White (85%), 9 (9%) as Asian, 3 (3%) as multiple races, 1 as African American or Black (1%), and 2 (2%) participants did not provide their race. One (1%) participant identified their ethnicity as Hispanic or Latine. Participants reported a range of ages, with 25-29 years being the most frequent category (n=24, 24%) and 30-34 (n=18, 18%) being the second most frequent category. No participants reported being age 65 or older. There were participants from all six NSGC regions, with Region 4 (AR, IA, IL, IN, KS, MI, MN, MO, ND, NE, OH, OK, SD, WI, Ontario) being the most frequently reported (n=32, 31%) and Region 2 ((DC, DE, MD, NJ, NY, PA, VA, WV, PR, VI, Quebec) being the second most frequently reported (n=24, 24%).

Professionally, participants had a variety of experience levels ranging from less than one year of experience to 30-34 years. The most common categories were 1-4 years (n=36, 35%) and 5-9 years (20, 20%). Seventy-four (73%) participants were in direct patient care positions, 23 (23%) were in mixed patient care positions, and 5 (5%) were in non-direct patient care positions. Participants worked in a variety of workplace settings. Forty-seven (46%) worked in an academic medical center, 21 (21%) in a public or community-based hospital, 13 (13%) in a private hospital or medical facility, 8 (8%) in a commercial laboratory, 1 (1%) in a non-hospital not-for-profit organization, 1 (1%) in private practice, and 11 (11%) in multiple settings.

### Genetic test ordering provider status

Ninety-three respondents (93%) provided information on their ordering status, and their demographic information is shown in Table 1. The majority of participants (n=56, 55%) placed orders but were not listed as the ordering provider of record. Twenty-seven (26%) respondents placed and signed genetic testing orders and were the provider of record. For the remaining respondents, 4 (4%) could not place or sign orders, 6 (5.9%) did not order in their role, 5 (5%) were not in a direct patient care role, and 4 (4%) did not respond.

**Table 1.**
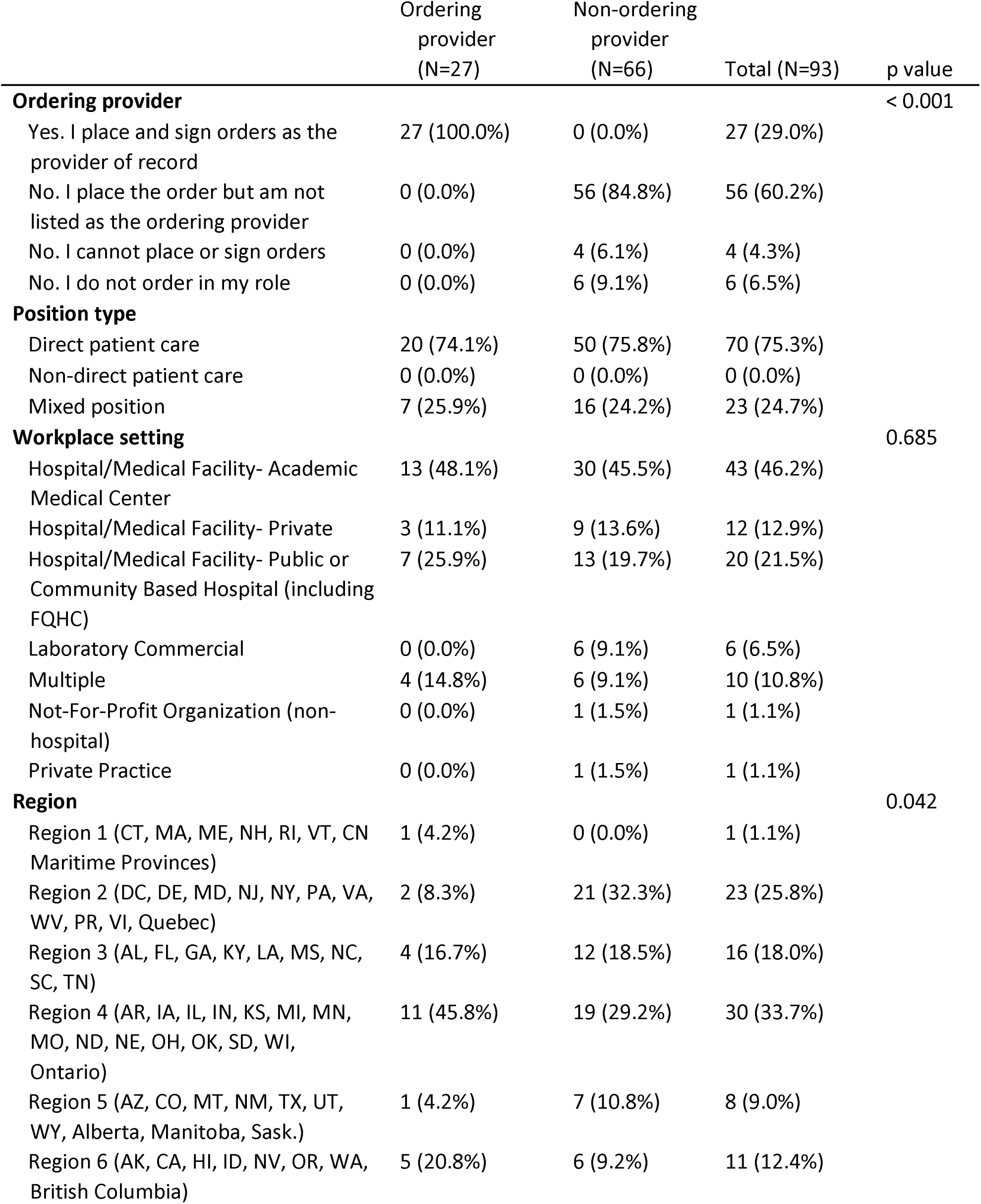
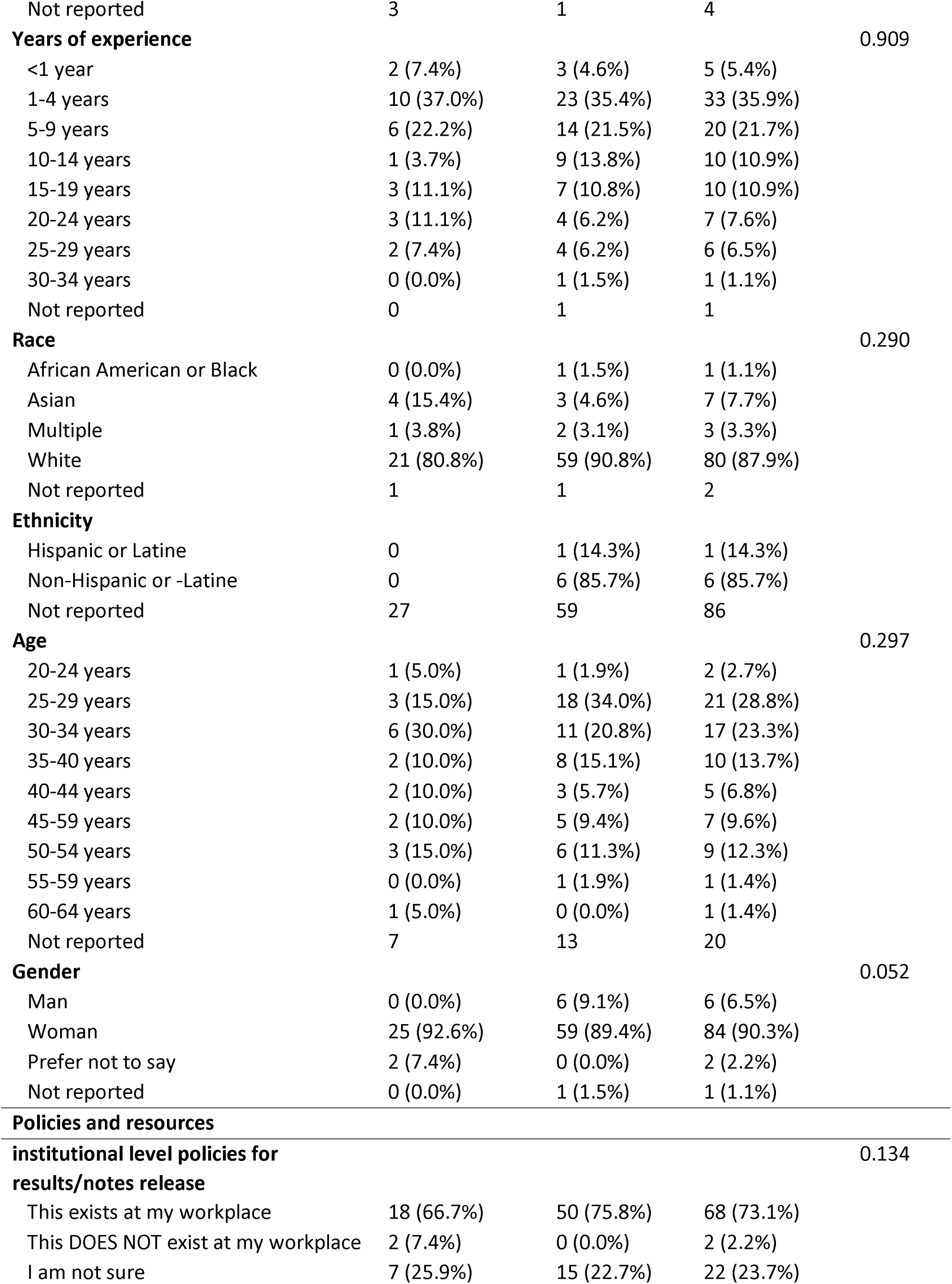
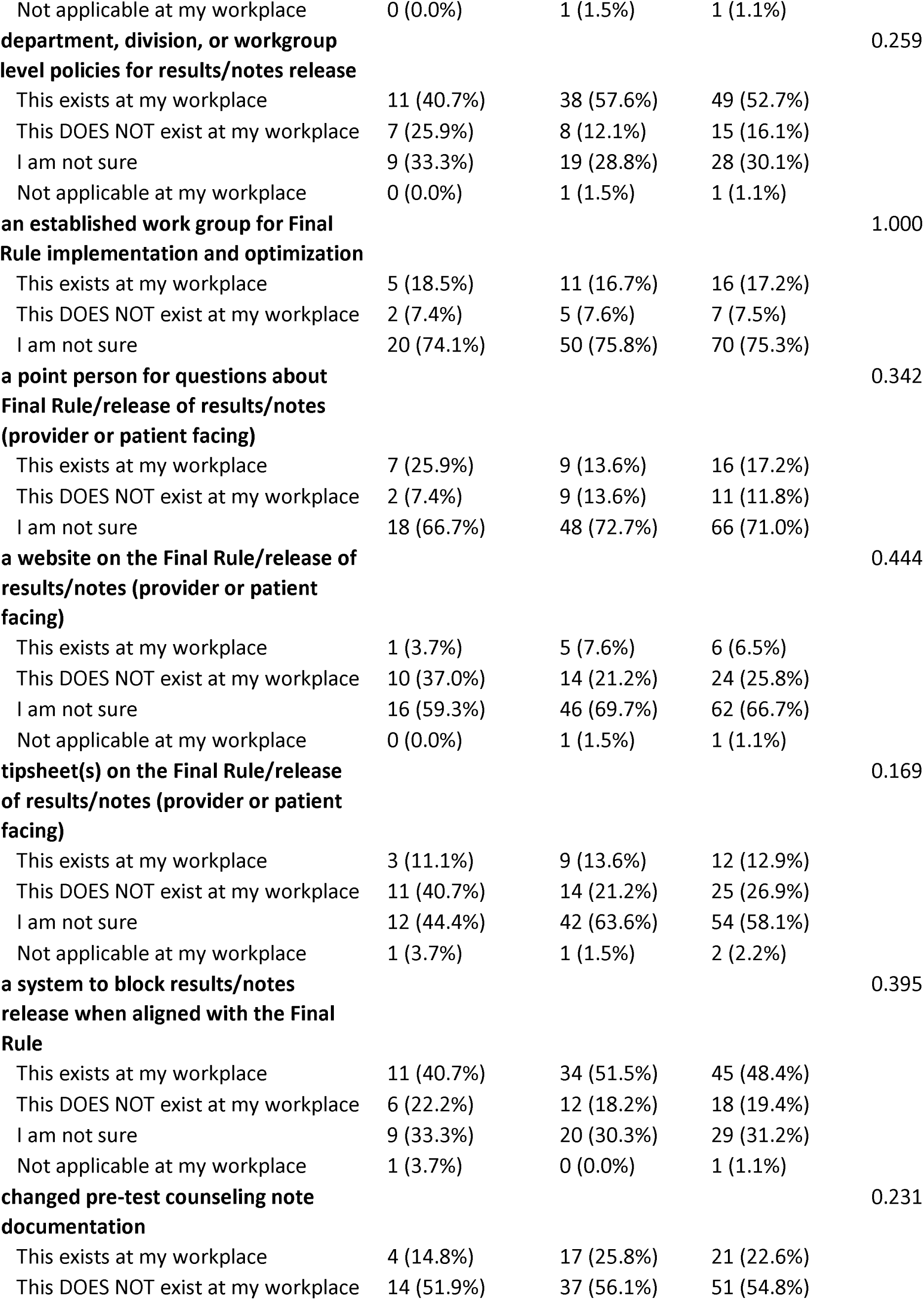
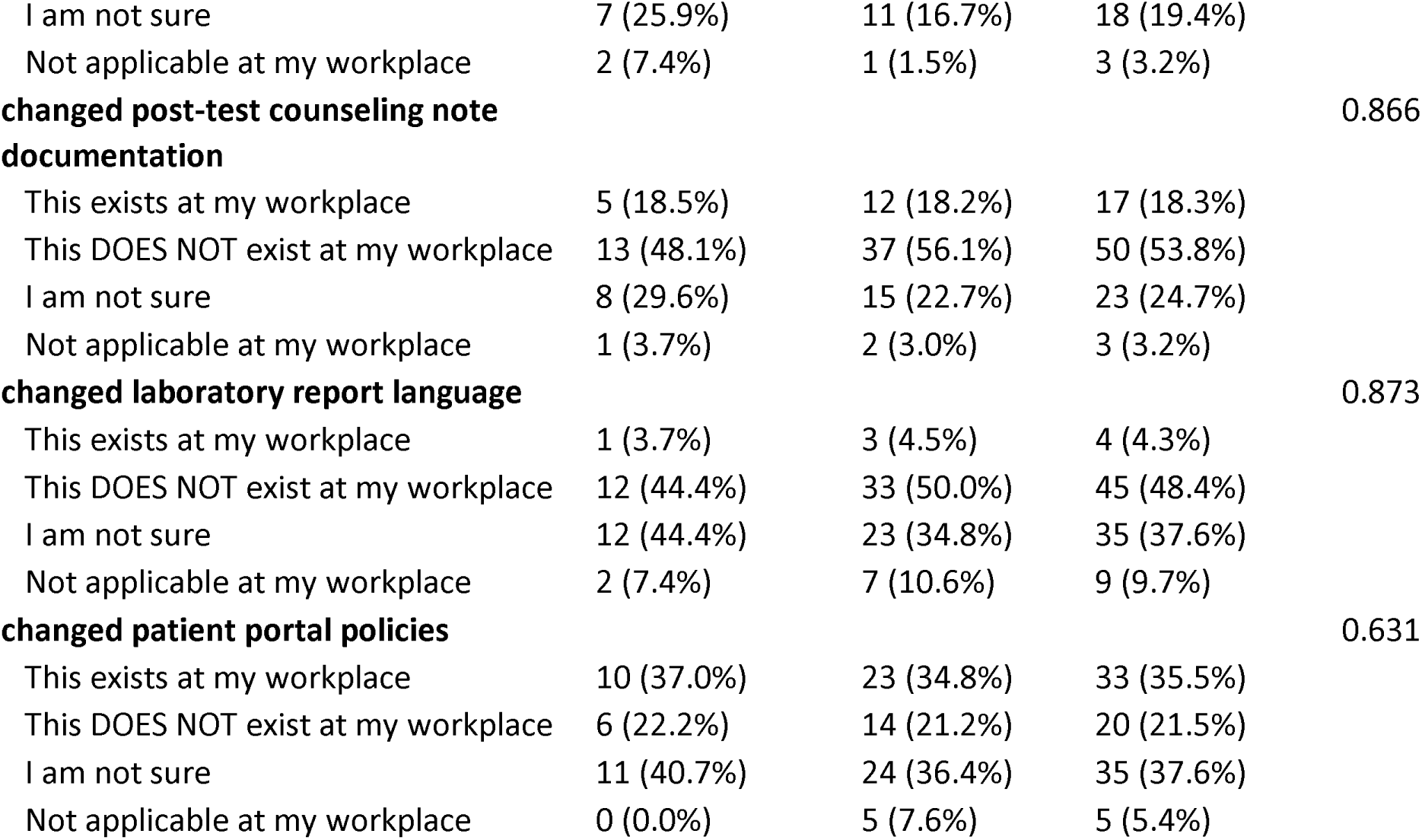
Comparison of ordering and non-ordering providers’ personal characteristics, workplace characteristics, and workplace policies related to the Final Rule.

To further explore how being an ordering versus non-ordering provider may have influenced the perceived impact of the Final Rule, we compared ordering providers (n=27) to all categories of patient-facing non-ordering providers (n=66) (Table 1). To facilitate statistical comparison, the three response options indicating a policy change was made were combined (“This exists and a GC was involved,” “This exists and a GC was NOT involved,” and “This exists and I don’t know if a GC was involved.”) The disaggregated responses are included in Supplemental Table 1. As shown in Table 1, the only statistically significant difference between the ordering and non-ordering providers was the geographic region (P=0.042); the largest proportion of ordering respondents reported living in Region 4.

Figures 1, 2, and 3 and Supplemental Table 2 compare ordering and non-ordering providers’ perceived workflow impact (Figure 1), harms (Figure 2), and benefits (Figure 3). While there were no statistically significant differences between ordering and non-ordering providers’ responses to the workflow impact items, there were statistically significant differences in responses for all three items pertaining to perceived patient harms (P values <0.05) and for one item pertaining to perceived patient benefits (P=0.007).

**Figure 1.**
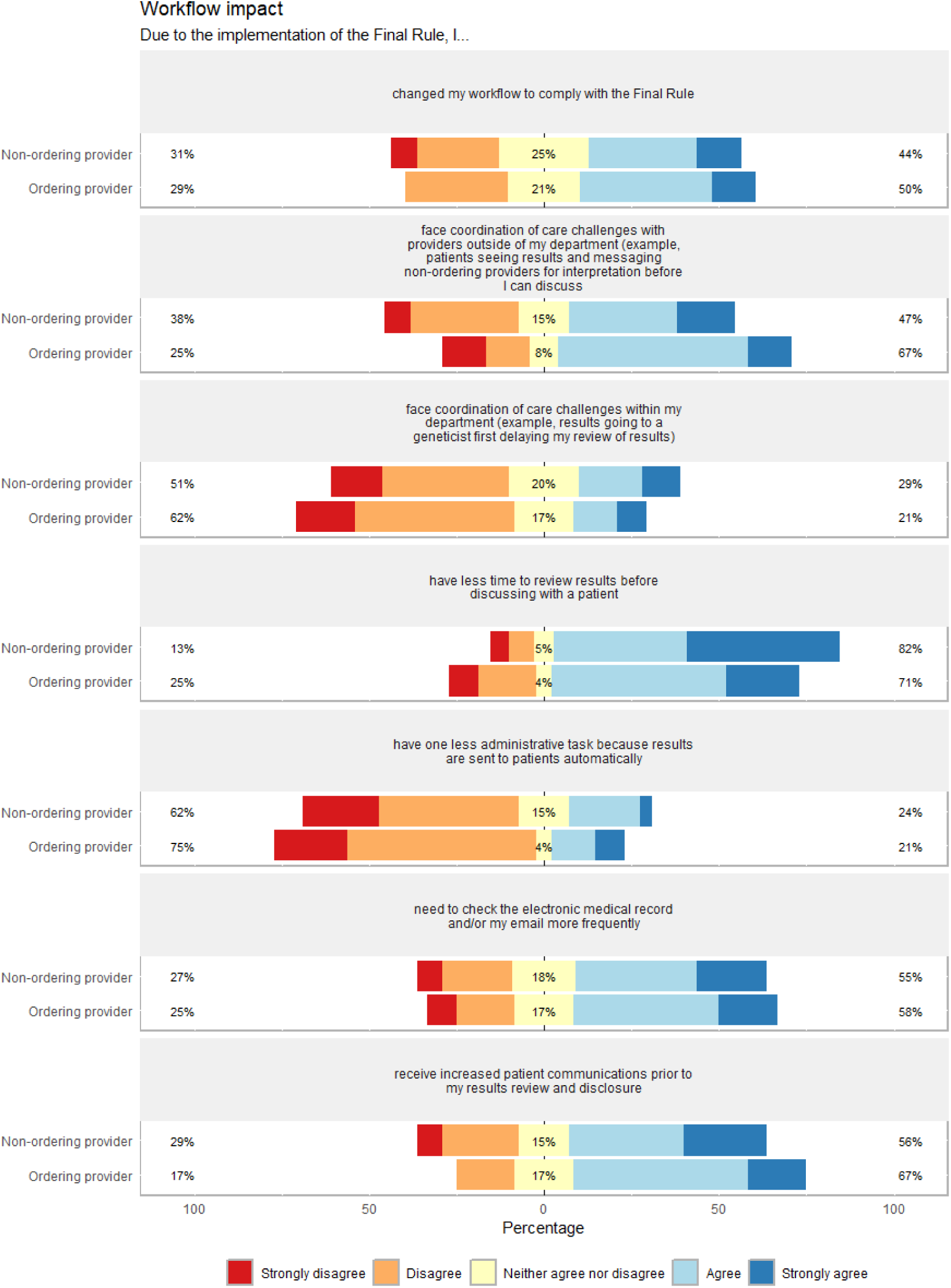
Genetic counselor-perceived workflow impacts of the Final Rule. Results are stratified by ordering provider status (if genetic counselors were the ordering provider for genetic tests ordered at their institution). Percentages on the left side (red and orange) reflect participants who strongly disagree (red) or disagree (orange) with the statement. Percentages on the right side reflect participants who agree (light blue) or strongly agree (dark blue). Percentages in the middle (yellow) of the chart reflect participants who neither agree nor disagree.

**Figure 2.**
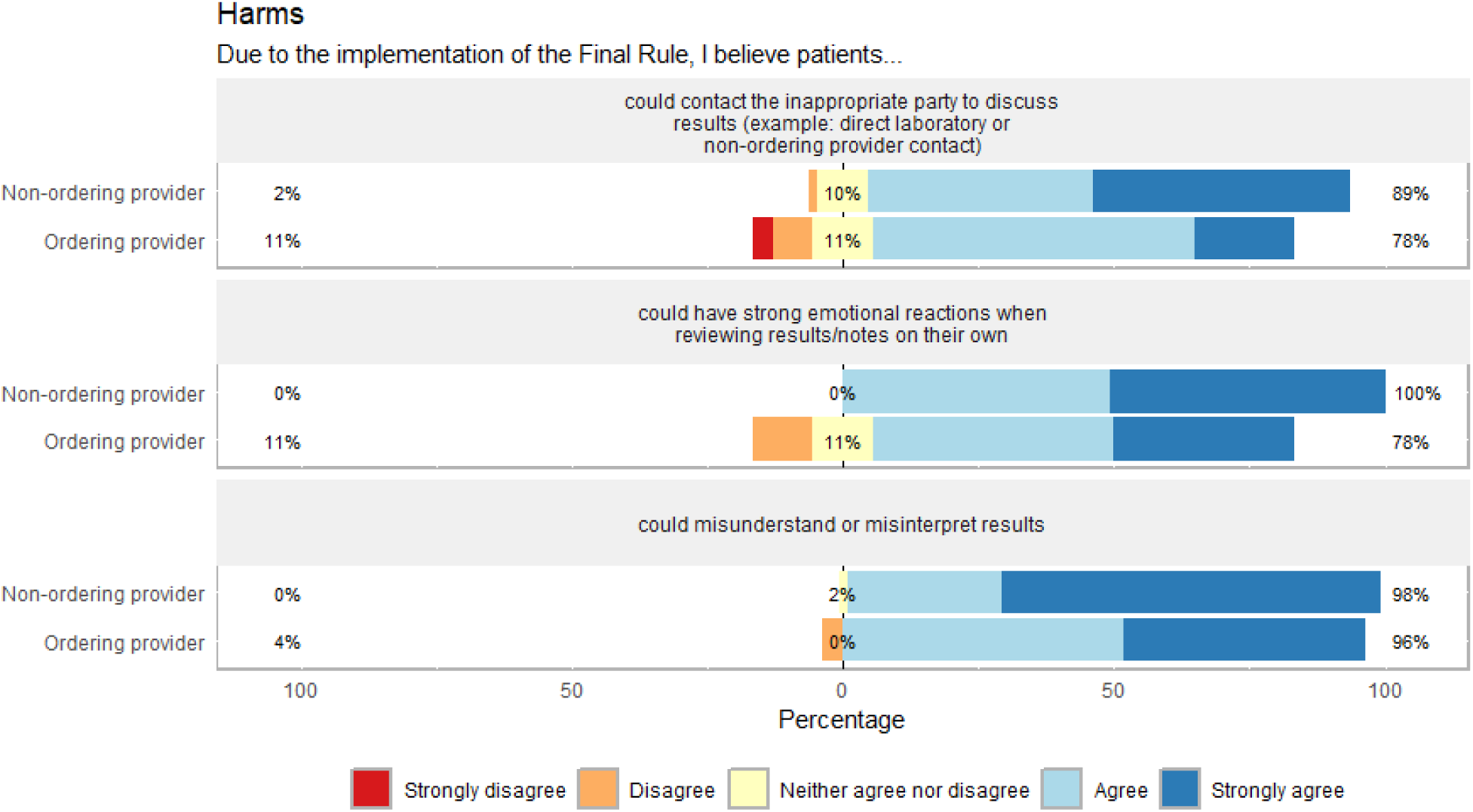
Genetic counselor-perceived patient harms due to the Final Rule, stratified by ordering provider status (if genetic counselors were the ordering provider for genetic tests ordered at their institution). Percentages on the left side reflect participants who strongly disagree (red) or disagree (orange) with the statement. Percentages on the right side reflect participants who agree (light blue) or strongly agree (dark blue). Percentages in the middle of the chart reflect participants who neither agree nor disagree (yellow).

**Figure 3.**
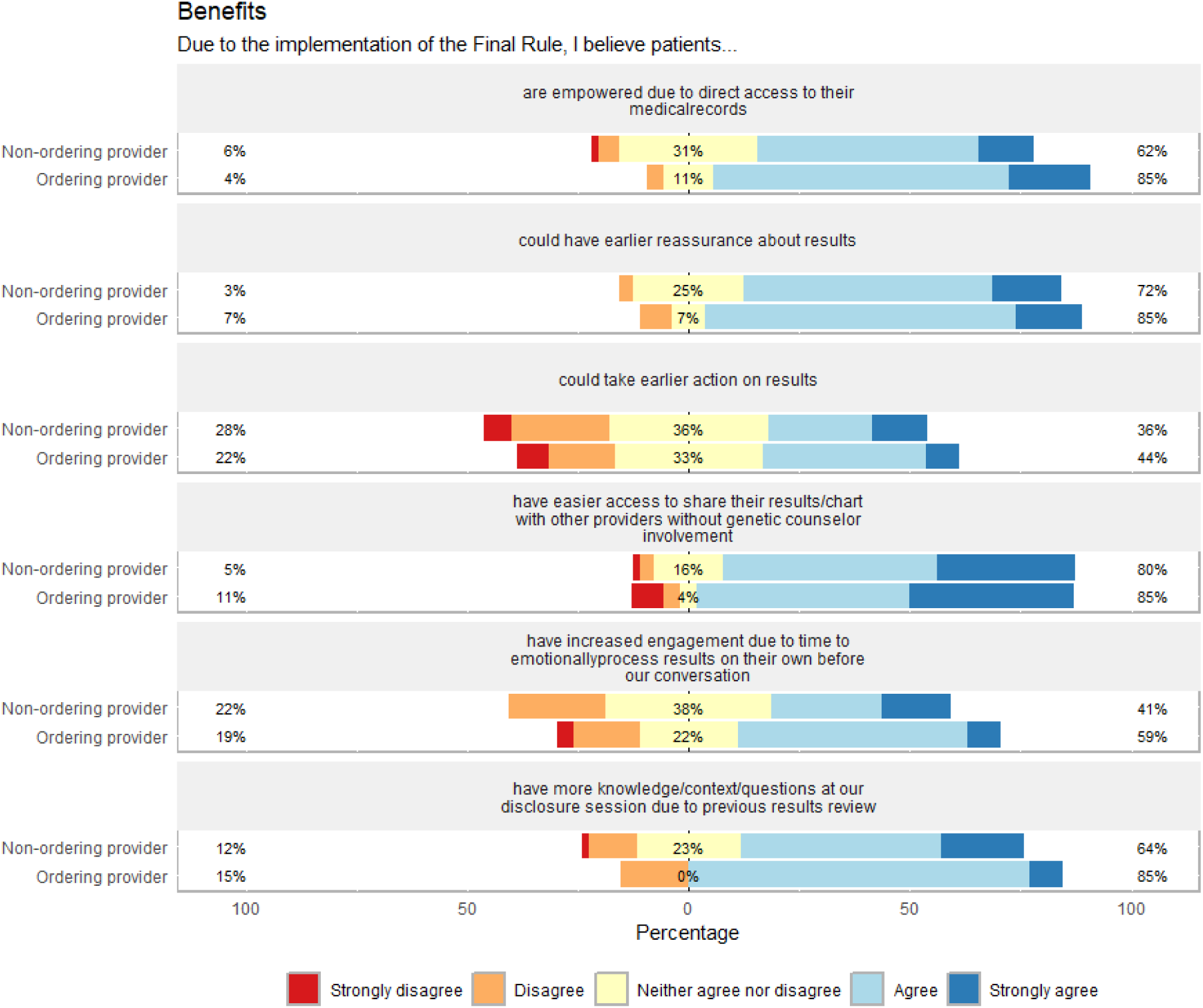
Genetic counselor-perceived patient benefits due to the Final Rule, stratified by ordering provider status (if genetic counselors were the ordering provider for genetic tests ordered at their institution). Percentages on the left side reflect participants who strongly disagree (red) or disagree (orange) with the statement. Percentages on the right side reflect participants who agree (light blue) or strongly agree (dark blue). Percentages in the middle of the chart reflect participants who neither agree nor disagree (yellow).

Non-ordering providers were more likely to “Strongly agree” with all three statements pertaining to patient harms. For the item “patients could have strong emotional reactions when reviewing results/notes on their own,” 51% of non-ordering providers selected “Strongly agree” compared to 33% of ordering providers (p=0.002). For the item “patients could misunderstand or misinterpret results,” 70% of non-ordering providers selected “Strongly agree” compared to 44% of ordering providers (P=0.025). For the item “patients could contact the inappropriate party to discuss results (example: direct laboratory or non-ordering provider contact),” 48% of non-ordering providers selected “Strongly agree” compared to 19% of ordering providers (P=0.023).

For the statement of benefit “patients have more knowledge/context/questions at our disclosure session due to previous results review,” responses were significantly different (P=0.007). Sixty-four percent of non-ordering providers agreed or strongly agreed compared to 81% of ordering providers.

### Qualitative responses to open-ended questions

Forty-nine (48%) participants provided at least one substantive response to at least one of the four optional open-ended survey questions. Numerical counts in this section refer to the number of times that the qualitative code was applied across all responses. The quotes presented here are ones which we judged to clearly illustrate the relevant category and provided additional experiential insight about Final Rule implementation that extended beyond the quantitative survey responses.

#### Negative impact

Responses to the “Negative impact” question (n=37) described negative impacts to both patients and providers.

##### Patient impact: Emotions

Sixteen responses mentioned that patients experienced distress related to direct release of results. This encompassed a range of levels of distress, with several examples of results in which families’ reactions were described as extreme, using terms such as “terrified” and “panic” (n=7):

> “Recently had a positive result released directly to the patient who called me in a panic because she did not understand the result…”

Respondents also mentioned a range of other patient reactions, including anxiety (n=4), being surprised or shocked (n=2), and anger or offense related to how results were disclosed via portal (n=2). Two respondents mentioned instances when Final Rule changes contributed to feelings of grief or regret:

> “I’ve had people find out fetal sex following a loss or termination that didn’t ever want that information…Learning the sex can complicate grieving and lead to unnecessary emotional distress for those that didn’t want to [know] fetal sex following a loss or termination.”

##### Patient impact: Misinterpretation

In two instances, respondents recounted situations in which misinterpreting results prior to counseling led families to begin to pursue inappropriate medical care:

> “Another family found out about a VUS [variant of uncertain significance] in the *WT1* gene. They then transfered [sic] care to a different hospital and told them that the patient had deny’s drash [sic] and patient almost underwent a radical nephrectomy before I was able to talk with parents/their new provider.”

There were additional examples of patients misunderstanding results prior to post-test counseling. Twelve respondents recounted instances of patients reading normal, uncertain, carrier, or screening results as diagnostic:

> “I have had patients who misinterpreted the reports who were leaning toward termination before I explained what the report actually meant and what was actually going on. For many of our patients, the Final Rule does not impact them in a meaningful way. But for a subset of patients, the outcome of the Final Rule is psychological torture.”

Conversely, three respondents provided examples of positive results being interpreted as normal:

> “I had one case where a patient result was released but they had only viewed 1 page (just *BRCA*) which was negative. They missed the full panel which was a few pages in, which had a pathogenic variant in another gene.”

Four respondents reported that their patients sought information online after receiving results directly before speaking with a clinician, potentially contributing to distress and misunderstanding.

### Patient impact: Timing of automatic release

Respondents were also concerned about the possibility of patients or families receiving potentially high-impact news through the impersonal medium of a patient portal (n=6) or of results being released at night or on a weekend when patients did not have immediate access to a clinician to help interpret the results (n=8). Participants emphasized both the emotional impact and the potential for misinterpretation when patients receive results when unable to contact a clinician:

> “An exome result was released after work on Friday revealing a likely pathogenic variant in 2 AR [autosomal recessive] genes and a pathogenic variant in a third AR gene. The family was terrified that their children had all three rare genetic conditions and were in tears when I reached them on Monday and said they hadn’t slept all weekend after seeing their child had a life limiting condition (which they did NOT have).”

> “I have a prenatal patient who had a fetal MRI on a Friday morning. The report was signed and released Friday end of day and that’s how she found out her baby has lissencephaly… How devastating is that? She had to sit with that information for an entire weekend with no one to contact.”

#### Provider impact: Workflow challenges

Perceived negative impacts of the Final Rule also included several workflow challenges. Six respondents described instances in which patients received results and contacted providers before the providers were able to review the results. Automatic release of results also led to a perceived increase in urgent requests from patients that negatively impacted genetic counselors’ other clinical duties and workflows (n=5):

> “One of my patients found out she was positive for both a *BRCA*1 pathogenic variant and a *BRCA2* pathogenic variant, while I was in the middle of clinic. I fortunately had a no show during that time, but given the complex results, I felt the need to call the patient immediately. It is tough with conversations about complex results that may take up to an hour and how that can impact the rest of my day’s agenda when patients randomly find out results.”

### Positive impact

Thirty-four (33%) respondents reported at least one positive impact of the Final Rule, including benefits to both patients and clinicians.

#### Patient impact: Affective benefits

Respondents frequently reported examples of patient benefits in affective terms that emphasized positive changes to patients’ preparedness, reassurance, and empowerment. Eight respondents noted that patients were more prepared for their genetic counseling visit when they had access to the results via automatic release:

> “It does allow processing time for those that understand their results and receive them in advance. For those patients it can make our initial interaction following the result more productive.”

Seven respondents discussed earlier patient reassurance:

> “automatic release benefits my most anxious patients. due to work hours and clinic schedules, I cannot always immediately call out results. patients whose anxiety is relieved by simply having a result (good or bad news) can receive a result as soon as it is available rather than as soon as I am available.”

Two respondents also mentioned that patients were more empowered in their care as a result of the Final Rule:

> “It empowers my patients to act as partners in their own medical care - they know that I am watching results, but that they can watch for results too. It allows families to better chose [sic] where and when they want to receive results, outside of the typical medical model of results disclosure appointments.”

#### Patient impact: Instrumental benefits

Respondents also described instrumental benefits to patients. These included: patients receiving results sooner (n=6), ease of accessing results and care (n=5), ease of sharing information and results with family members (n=4), and reducing the possibility that test results are never reported or shared with the patient (n=3).

#### Provider impact: Workflow efficiency

The most commonly reported perceived benefit to clinicians was that changes related to the Final Rule helped them work more efficiently (n=11). Examples of improvements included automatic results release and decreased need to update patients while results are pending:

> “…they can sign up for the patient portal ahead of time so they’ll be notified when their results are available, instead of calling me every day to ask if the result is out yet. It has made my life easier in that respect.”

### Impact of patient and workplace characteristics

Thirty (29%) respondents to the free response prompts reported at least one patient characteristic that they felt influenced patients’ experiences of risks or benefits related to Final Rule changes. The most frequent characteristic that respondents mentioned was limited access to technology or to the patient portal (n=11). This sentiment was also present in those that felt there was the potential to foster distrust (n=1) and widen existing health disparities by reducing benefits for patients with limited access to portals and accentuating potential harms among patients who are less equipped to make sense of automatically-released results (n=5). Respondents also frequently expressed concerns about the impact on patients with limited English proficiency (n=7), patients with low health literacy or education (n=6), and patients from low socioeconomic or otherwise underserved backgrounds (n=6):

> “The medical record is not translated into other languages so this rule is limited for non english [sic] speakers or those that can read in english.”

> “Most of my patients choose not to use the EHR [electronic health record] because they either have low computer literacy or do not have reliable internet access. So the new rules only impact a subset of my patient population. Honestly the people who don’t use our EHR would probably be more likely to experience the negative side of the rules (fear,misinterpretation,etc [sic]).”

In addition to these access-related challenges, respondents also mentioned individual patients’ mental health (n=3), age (n=2), disabilities (n=1), and degree of personal interest in genetics (n=1) as factors that influenced patients’ experiences of automatic release of results and other Final Rule changes.

Several respondents discussed practicing in settings where patients came from more privileged backgrounds, including high socioeconomic status, health literacy, education, and technology literacy. While some respondents described these features as being protective against potential negative impacts of the Final Rule, two recognized that the Final Rule posed challenges even to relatively advantaged patients:

> “I work with a largely well educated population that is very tech savvy. But even this population has struggles with this rule.”

### Practice changes

#### Pre-test counseling

Thirty-three (32%) qualitative respondents provided at least one example of a practice change made in response to the Final Rule, with more than half mentioning changes to pre-test counseling. Eighteen respondents described informing patients during pre-test counseling that they may receive results before the provider is able to access or review them:

> “I now have to explicitly explain to patients that there are two timeframes: the turnaround time of the test and the time it takes to interpret the results in clinical context. I also have to remind patients that I will call them after both of these things have taken place so they may see their result a few weeks before hearing from me.”

Additional changes to pre-test counseling included counseling patients to consider waiting to access results that are released automatically (n=6), setting expectations about how results will arrive (n=4), and discussing the option of consenting to block automatic release of results (n=1).

#### Return of results

Regarding return of results, four respondents described feeling more pressure to review and call out results quickly. However, two respondents reported that automatically released results reduced the administrative burden and urgency of contacting patients.

#### Clinical documentation

Qualtiative respondents provided seven examples of changes to documentation practices. Three respondents mentioned increased awareness and concern about documenting patients’ psychosocial concerns or sensitive information in clinical notes that patients can access:

> “It’s difficult to comment on the psychosocial state of a patient, so many times I leave it out, which is a disservice to other providers. This is especially true when there is concerning behavior. Patients have asked me to leave out details like termination of pregnancies and even family history details.”

Respondents also reported documenting that results would be immediately released to patients in the pre-test note (n=2), writing a separate patient-friendly note and results letter that are released on the same day as the result (n=1), and increasing the patient-friendliness of clinical documentation (n=1). However, this last respondent expressed concerns about the added administrative burden of this practice:

> “We were given a tip sheet on documentation to be more patient friendly. I think it’s good for us to think about how we write things and make language accessible. But you’re asking doctors to see 40 pts [patients] in a day and also think critically about their note? That’s burn out.”

#### Lab report documentation

Two responses described changes to lab genetic counselors’ practices. One discussed adding a disclaimer to lab reports to encourage patients to contact the ordering provider rather than the lab. Another reported changing their approach to writing result reports:

> “…the draft results reports I write are different now, because I know patients will see them in their entirety and possibly before the ordering provider.”

### Cross-cutting categories

We identified two recurring categories that were not linked to a specific survey question: Non-exceptionalism of genetic results and potential solutions and normative claims.

#### Non-exceptionalism of genetic results

Three respondents noted ways in which genetic results were not exceptional when compared to other kinds of medical results. Two responses emphasized that distress or confusion upon receiving results is not unique to genetics and that clinicians in pathology and other specialties manage similar reactions. included examples of patients’ distress or confusion after receiving non-genetic results such as imaging, pathology, or lab results. One respondent voiced support for applying similar standards to genetics and other types of medical results:

> “Patients receive all other results immediately - including other sensitive results such as dx [diagnosis] of cancer and pregnancies related tests. So having genetic tests be treated like other lab tests is a step in the right direction, in my opinion, as we continue to see genetics move into the mainstream of [care].”

#### Potential solutions and normative claims

Although none of the four open-response questions directly solicited solutions for how genetic services should be provided in the context of the Final Rule, four respondents offered suggestions (Supplemental Table 3). One respondent voiced concerns about the appropriateness of immediate release, especially for patients receiving high-impact results such as in adult neurology. Similarly, one respondent voiced general support for the Final Rule but proposed a short delay to mitigate the potential harms of immediate release. Another respondent recognized the utility of providing an immediate point of contact:

> “They need someone to call who gives immediate feedback when they have questions. Just enough info to relieve anxiety before their next visit with the ordering provider.”

One respondent called for increased attention to disparities and unequal distribution of benefits due to Final Rule changes:

> “I think there are some patient demographics who are more likely to use the benefits of the Final Rule than others so we should just be mindful of that in office policies regarding release of results to make sure those less likely to use the benefits still get their results.”

## Discussion

Both quantitative and qualitative responses indicated that genetic counselors perceived both positive and negative impacts on patients, providers, and workflow efficiency due to the Final Rule. Qualitative analysis of open-ended responses helped to triangulate and contextualize the quantitative results, raising new directions for future research.

### Patient harms and benefits

Regarding patient harms and benefits, there was good alignment between the quantitative survey and the open-ended responses. Both closed- and open-ended responses indicate that genetic counselors are concerned about the potential emotional impact on patients receiving results automatically and that they worry about patients misunderstanding their results. Examples from the open-ended responses indicate that these concerns are not merely theoretical but rooted in prior experience – in some cases weighing heavily on respondents.

Conversely, genetic counselors recognized a range of benefits to patients, with more than 50% of respondents agreeing or strongly agreeing with all survey questions that described potential patient benefits, except for the statement that patients “could take earlier action on results.” In the open-ended responses, respondents also recognized the additional benefits of patients being able to more easily share information with their family members and reducing the possibility of results being “lost” due to not being reported to the patient or not being incorporated into the medical record.

### Adapting to the Final Rule

Fewer than half of respondents (46%) reported changing their workflow to comply with the Final Rule. Synthesizing the quantitative and qualitative responses, most of these changes appear to either increase or disrupt work rather than reducing it, with more than 50% of respondents agreeing or strongly agreeing with statements related to coordination of care challenges, having less time to review results before discussing with the patient, needing to check the electronic medical record and/or email more frequently, and receiving increased patient communications prior to results disclosure. In the open-ended questions, respondents described additional workflow disruptions including changes to clinical documentation.

Several open-ended and close-ended survey responses did indicate positive, though perhaps minimal, changes to workflow efficiency. Several open-ended responses mentioned increased workflow efficiency due to automatic release of results, but only 23% of respondents agreed or strongly agreed that they now have one less administrative task because results are sent to patients automatically. This suggests that the procedure for releasing results either did not change due to the Final Rule for the majority of respondents and/or that respondents did not perceive automatic release as having a large impact on their workflow. Additionally, several open-ended responses discussed decreased urgency of disclosing normal results as a beneficial change in their workflow when they see that the patient has already viewed the report. However, it is unclear if this reduces genetic counselors’ overall workload or allows genetic counselors to better prioritize their time.

While the overall impact of the Final Rule on genetic counselors’ time and work efficiency is uncertain and likely depends on specific workplace characteristics, the possibility of adding to genetic counselors’ administrative burden should be taken seriously because of the potential implications for work satisfaction and burnout (Caleshu et al., 2022). Genetic counselors may therefore benefit from evaluating the impact of the Final Rule on their workplace efficiency and advocating for increased administrative support or more streamlined processes if the Final Rule has increased their administrative burden.

Although respondents discussed a variety of measures that they and their institution have taken to mitigate these potential harms, there may still be room for improvement. While more than 50% of respondents reported having institutional-and/or department-, division-, or workgroup-level policies for results and notes release, less than half of respondents were aware of having any of the other resources or policies (e.g., established work group, point person for questions, website resources, tip sheet) described in the survey. Perhaps most notable is that only 48% of respondents were aware of a system to block results/notes release that is aligned with the Final Rule, with 31% of respondents being unsure of whether such a system existed at their institution (Supplemental Table 1). The relatively high levels of genetic counselors reporting that they are not sure if a policy or resource exists suggests a need for better communication about Final Rule changes within institutions.

### Ordering vs. non-ordering providers

The comparisons of ordering and non-ordering providers were conducted to better understand the extent to which genetic counselors’ ability to place orders and directly receive genetic test results might influence their and their patients’ experiences of the Final Rule. While prior literature on clinicians’ experiences of the Final Rule has primarily focused on physicians, ordering provider status is one respect in which this literature may not fully translate to genetic counseling practice.

Non-ordering providers were more likely to strongly agree with statements about potential harms to patients than ordering providers, including concerns for patients’ emotional reaction, misunderstanding or misinterpreting results, and contacting the inappropriate party to discuss results. Non-ordering providers were less likely than ordering providers to agree or strongly agree with one statement conveying a perceived benefit to patients: “patients have more knowledge/context/questions at our disclosure session due to previous results review.” These findings raise the possibility that because non-ordering providers might receive results later than ordering providers, they are more concerned about longer delays between the patient receiving the results and having the opportunity to discuss them with a genetic counselor. This is consistent with some findings from the NICU setting – a setting that, like genetic counseling, tends to take a team-based approach. In this study, NICU staff members acknowledged greater possibility for patient misunderstanding with immediate release in a setting where the provider who coordinated testing may not be the first provider with whom a family chooses to speak (Sullivan et al., 2024).

In genetic counseling, patients receiving reports that list only the name of the ordering clinician may similarly be unclear about who to contact with questions about the report. This may be especially problematic for clinical models where the ordering provider does not meet the patient or where the ordering provider does not have genetics expertise. While additional research is needed to determine if non-ordering providers’ concerns reflect patient-perceived harms and benefits, ensuring that genetic counselors can directly receive result reports may help to mitigate some provider-perceived harms.

Possible models for this include having genetic counselors serve as ordering providers (if allowed by institutional and state policies) or by adding genetic counselors to orders as additional providers who receive notification at the same time as the ordering provider.

### Suggestions

Informed by organizational health literacy principles (*Health Literacy in Healthy People 2030 - Healthy People 2030 | Health.Gov*, n.d.) and drawing from the concerns, case examples, and practice changes described or endorsed by the survey respondents, we have compiled a preliminary list of suggestions for members of the genetic counseling community for navigating the Final Rule.

1. **Provide pre-test anticipatory guidance to patients about how results will be returned and discuss options for return of results.** Many survey respondents mentioned pre-test counseling as a way of mitigating potential harms from automatic return of results. Letting patients know that they could receive results before a clinician is prepared to discuss the results and encouraging them to wait to discuss the results with the appropriate clinician can help to reduce surprise, misunderstanding, and inappropriate care. During pre-test conversations, patients can also consider if they would prefer to delay seeing the result report – either by not opening the report when it is released to the patient portal or by consenting to temporarily block automatic release of results (if institutional policy and electronic health record systems allow). Patient preferences and requests to block automatic release of results should be clearly documented.
2. **Consider legally compliant and patient-centered procedures to blocking automatic release of results.** Both closed- and open-ended survey responses indicate that genetic counselors are concerned about automatic return of results causing emotional distress and potential misunderstandings before they have the opportunity to discuss with a provider. Prior research exploring patient preferences suggests patient support for the ability to opt out of automatically released results (Bruno et al., 2022). While a temporary and consensual block would potentially mitigate many genetic counselors’ concerns about patient misunderstandings and emotional distress, approximately half of respondents either did not have a way to block automatic release of results and clinic notes or they were not aware of how to do so. Institutions and genetic services can potentially examine how procedures were implemented at other institutions to shape their own legally compliant and patient-centered procedures to offer patients the option to temporarily block automatic release of results.
3. **Implement procedures to keep genetic counselors “in the loop” during test ordering and disclosure.** Without clear policies and communication channels, the Final Rule has the potential to leave genetic counselors temporarily “out of the loop” when results are returned and the genetic counselor is not the ordering provider, resulting in delayed outreach to patients. Genetic counselors who are not ordering providers may benefit from the capability for multiple providers (including non-ordering genetic counselors) to automatically receive test results if such a capability does not already exist.
4. **Adopt a universal precautions approach to mitigate health disparities.** A frequent negative outcome reported by respondents was increased confusion, distress, or misinterpretation of results, which can be exacerbated by limited medical literacy or English literacy. A universal precautions approach involves adopting procedures that assume that every patient is at risk of not understanding or being able to act appropriately on health information (Paasche-Orlow et al., 2006). Moreover, the environment, not the patient, needs to change (*Health Literacy in Healthy People 2030 - Healthy People 2030 | Health.Gov*, n.d.). Adopting a universal precautions approach to the Final Rule would involve implementing standard pre-test, reporting, and return of results procedures that are designed for patients who are at the highest risk for misunderstanding or experiencing emotional distress from genetic information. These procedures should be tailored to the specific clinical context but might include:

- Routine incorporation of accessible educational materials about genetic testing into genetic reports and return of results documentation
- Discussing what patients should do if they receive results that are confusing or distressing
- Offering the option to temporarily block automatic return of results
- Using patient-friendly language during clinical documentation
- Striving for educational and results resources that are translated into many languages

testing laboratories can further support this by following best practices for health literacy in the patient-friendly design and content of genetic test reports. Although a universal precautions approach does not necessarily mitigate disparities related to unequal benefit from the Final Rule, it is an important step toward preventing unequal distribution of patient benefits and harms.
5. **Educate non-genetics ordering providers about Final Rule considerations for pre-test counseling, ordering, and return of genetic results.** Genetic testing is frequently ordered by non-genetics providers in settings such as cancer (McCuaig et al., 2018). Non-genetics providers may be less familiar with the psychosocial impact of genetic results and with the legal standards that allow patients to consent to block automatic release of genetic results. Therefore, there may be a need for outreach to non-genetics ordering providers to formulate patient-centered return of results procedures and to inform them of best practices for pre-test counseling, including discussing whether patients wish to consent to block automatic release of results.
6. **Recognize that not all patients will have access to their results after Final Rule implementation and develop alternative ways for patients to easily access their results.** Respondents emphasized that limited computer literacy, English literacy, and access to the Internet can impact patients’ ability to readily access their results as described by the Final Rule. Institutions should work with their local patient communities to create alternatives for receiving results outside a patient portal that are simple, free, and have instructions provided in many languages.

These suggestions have important limitations, including that they are based on limited and potentially non-representative evidence from a national survey. Moreover, they do not directly incorporate patient perspectives. Policies should be tailored to the specific needs of the local institution and patients.

Consultation with the relevant institutional authorities, providers, and patient representatives can help ensure that policy or practice changes are legally compliant and suitable for the institution and patient population.

### Directions for future research

This study examined genetic counselors’ perceptions of patient harms and benefits and did not assess patients’ direct report of harms or benefits. Future research examining patient experiences of Final Rule implementation can help to determine the extent to which providers’ attempts to mitigate patient harms are aligned with patient needs and preferences.

Although genetic counselors in non-direct patient care positions were eligible to complete the survey, there was relatively low response from genetic counselors in laboratory and other non-patient facing roles. Although the survey responses provide some limited insight into how laboratory genetic counselors have responded to the Final Rule, future studies can build on this to address how the Final Rule affects genetic counselors in non-patient facing roles and to identify possible opportunities for laboratory-based genetic counselors to help mitigate the potential for patient misunderstanding of automatically-released reports and confusion about the correct points of contact.

### Limitations

This survey has a number of limitations related to both internal and external validity. Due to the lack of available validated measures pertaining to Final Rule implementation, we created novel survey items to assess genetic counselors’ attitudes and workplace policies that were informed by prior exploratory work (Morris, 2022); however these measures did not undergo any prior psychometric evaluation and should be interpreted with caution. Regarding the qualitative findings, a major limitation is that the open-ended survey questions did not allow for additional probing to explain or elaborate on responses. Moreover, thematic saturation was not reached, with new categories continuing to emerge throughout the coding process. But while there were limitations to the qualitative analysis that could be performed, it did offer additional value to our quantitative analysis by providing context through which to interpret the findings and expanding our interpretation of the Final Rule’s impacts beyond the pre-specified quantitative variables included in the survey.

A limitation of the analysis of ordering and non-ordering providers is that we had to combine different types of non-ordering providers to facilitate statistical analysis. Due to a survey error, we were unable to capture which non-ordering providers did and did not receive direct notifications of test results and therefore could not compare these groups. Therefore, we cannot determine the extent to which notification practices may account for the differences between ordering and non-ordering providers’ perceived impact of the Final Rule. In addition, although the only difference in personal or workplace characteristics between ordering and non-ordering providers was in the geographic region – which may reflect differences in state laws about whether genetic counselors are authorized to order genetic testing – we cannot rule out the possibility of other confounding influences such as electronic health record capabilities and amount of administrative support.

Findings from this survey may not be generalizable to all U.S. genetic counselors due to the likelihood of this being a non-representative sample. Since recruitment methods included promoting the survey following a plenary presentation at an NSGC Annual Conference and use of NSGC listserv emails, genetic counselors who are more actively engaged with NSGC are likely to be overrepresented. It is also possible that multiple genetic counselors from the same institution may have responded, leading to non-independent responses for survey items about workplace characteristics and policies. The survey responses are subject to self-selection bias, perhaps with genetic counselors with strong opinions about the Final Rule implementation being more likely to complete the survey. Additionally, 101 participants started but did not complete the survey. While we hypothesize that the high rate of survey non-completion may have been due to the fact that conference attendees likely needed to move on to attend subsequent conference sessions after the plenary presentation, we cannot determine if respondents who started but did not complete the survey may have differed from those who completed the survey.

### Practice Implications

The survey responses illustrate a diversity of approaches to handling the Final Rule implementation at institutional, clinic, and individual levels. While implementation approaches should be tailored to the specific needs of individual clinics, genetic counselors who are struggling with aspects of Final Rule implementation may benefit from learning how other genetic counselors and institutions are creating solutions to similar challenges.

## Conclusion

The implementation of the Final Rule not only changes paradigms of information sharing across medical specialties but also raises specific issues for genetic services due to the psychosocial and informational complexities of genetic information. Our data show that genetic counselors have a variety of concerns and perceived benefits related to the Final Rule’s implementation. Moreover, genetic counselors’ ability to order and receive results from genetic testing may moderate concerns about potential negative impacts on patient care. These quantitative and qualitative findings suggest possible strategies to mitigate negative impacts on patient care, but further research is needed to understand the extent to which these mitigation strategies meet patients’ needs and promote efficient use of genetic counseling resources.

## Ethics approval statement

This study was approved as exempt by the Mayo Clinic Institutional Review Board (23-007555).

## Data availability statement

The data that support the findings of this study are available on request from the corresponding author.

## Supporting information

survey code book

## Acknowledgments

The authors thank the study participants.

