## Supplemental tables for "Navigating the Impact of the 21st Century Cures Act Final Rule: A National Cross-Sectional Survey of U.S. Genetic Counselors"

### Supplemental data

Supplemental Table 1. Disaggregated table of workplace policies comparing ordering and non-ordering providers

|  | Ordering provider (N=27) | Non-ordering provider (N=66) | Total (N=93) | p value (Fisher’s exact test) |
| --- | --- | --- | --- | --- |
| **institutional level policies for results/notes release** |  |  |  | 0.115 |
| This exists and a GC was involved | 3 (11.1%) | 6 (9.1%) | 9 (9.7%) |  |
| This exists and a GC was NOT involved | 13 (48.1%) | 27 (40.9%) | 40 (43.0%) |  |
| This exists and I don’t know if a GC was involved | 2 (7.4%) | 17 (25.8%) | 19 (20.4%) |  |
| This DOES NOT exist | 2 (7.4%) | 0 (0.0%) | 2 (2.2%) |  |
| I am not sure | 7 (25.9%) | 15 (22.7%) | 22 (23.7%) |  |
| Not applicable at my workplace | 0 (0.0%) | 1 (1.5%) | 1 (1.1%) |  |
| **department, division, or workgroup level policies for results/notes release** |  |  |  | 0.226 |
| This exists and a GC was involved | 6 (22.2%) | 19 (28.8%) | 25 (26.9%) |  |
| This exists and a GC was NOT involved | 5 (18.5%) | 10 (15.2%) | 15 (16.1%) |  |
| This exists and I don’t know if a GC was involved | 0 (0.0%) | 9 (13.6%) | 9 (9.7%) |  |
| This DOES NOT exist | 7 (25.9%) | 8 (12.1%) | 15 (16.1%) |  |
| I am not sure | 9 (33.3%) | 19 (28.8%) | 28 (30.1%) |  |
| Not applicable at my workplace | 0 (0.0%) | 1 (1.5%) | 1 (1.1%) |  |
| **an established work group for Final Rule implementation and optimization** |  |  |  |  |
| This exists and a GC was involved | 1 (3.7%) | 4 (6.1%) | 5 (5.4%) |  |
| This exists and a GC was NOT involved | 2 (7.4%) | 6 (9.1%) | 8 (8.6%) |  |
| This exists and I don’t know if a GC was involved | 2 (7.4%) | 1 (1.5%) | 3 (3.2%) |  |
| This DOES NOT exist | 2 (7.4%) | 5 (7.6%) | 7 (7.5%) |  |
| I am not sure | 20 (74.1%) | 50 (75.8%) | 70 (75.3%) |  |
| Not applicable at my workplace | 0 (0.0%) | 0 (0.0%) | 0 (0.0%) |  |
| **a point person for questions about Final Rule/release of results/notes (provider or patient facing)** |  |  |  |  |
| This exists and a GC was involved | 3 (11.1%) | 3 (4.5%) | 6 (6.5%) |  |
| This exists and a GC was NOT involved | 3 (11.1%) | 3 (4.5%) | 6 (6.5%) |  |
| This exists and I don’t know if a GC was involved | 1 (3.7%) | 3 (4.5%) | 4 (4.3%) |  |
| This DOES NOT exist | 2 (7.4%) | 9 (13.6%) | 11 (11.8%) |  |
| I am not sure | 18 (66.7%) | 48 (72.7%) | 66 (71.0%) |  |
| Not applicable at my workplace | 0 (0.0%) | 0 (0.0%) | 0 (0.0%) |  |
| **a website on the Final Rule/release of results/notes (provider or patient facing)** |  |  |  | 0.215 |
| This exists and a GC was involved | 1 (3.7%) | 0 (0.0%) | 1 (1.1%) |  |
| This exists and a GC was NOT involved | 0 (0.0%) | 4 (6.1%) | 4 (4.3%) |  |
| This exists and I don’t know if a GC was involved | 0 (0.0%) | 1 (1.5%) | 1 (1.1%) |  |
| This DOES NOT exist | 10 (37.0%) | 14 (21.2%) | 24 (25.8%) |  |
| I am not sure | 16 (59.3%) | 46 (69.7%) | 62 (66.7%) |  |
| Not applicable at my workplace | 0 (0.0%) | 1 (1.5%) | 1 (1.1%) |  |
| **tipsheet(s) on the Final Rule/release of results/notes (provider or patient facing)** |  |  |  | 0.340 |
| This exists and a GC was involved | 1 (3.7%) | 1 (1.5%) | 2 (2.2%) |  |
| This exists and a GC was NOT involved | 2 (7.4%) | 6 (9.1%) | 8 (8.6%) |  |
| This exists and I don’t know if a GC was involved | 0 (0.0%) | 2 (3.0%) | 2 (2.2%) |  |
| This DOES NOT exist | 11 (40.7%) | 14 (21.2%) | 25 (26.9%) |  |
| I am not sure | 12 (44.4%) | 42 (63.6%) | 54 (58.1%) |  |
| Not applicable at my workplace | 1 (3.7%) | 1 (1.5%) | 2 (2.2%) |  |
| **a system to block results/notes release when aligned with the Final Rule** |  |  |  | 0.411 |
| This exists and a GC was involved | 2 (7.4%) | 6 (9.1%) | 8 (8.6%) |  |
| This exists and a GC was NOT involved | 7 (25.9%) | 14 (21.2%) | 21 (22.6%) |  |
| This exists and I don’t know if a GC was involved | 2 (7.4%) | 14 (21.2%) | 16 (17.2%) |  |
| This DOES NOT exist | 6 (22.2%) | 12 (18.2%) | 18 (19.4%) |  |
| I am not sure | 9 (33.3%) | 20 (30.3%) | 29 (31.2%) |  |
| Not applicable at my workplace | 1 (3.7%) | 0 (0.0%) | 1 (1.1%) |  |
| **changed pre-test counseling note documentation** |  |  |  |  |
| This exists and a GC was involved | 4 (14.8%) | 15 (22.7%) | 19 (20.4%) |  |
| This exists and a GC was NOT involved | 0 (0.0%) | 0 (0.0%) | 0 (0.0%) |  |
| This exists and I don’t know if a GC was involved | 0 (0.0%) | 2 (3.0%) | 2 (2.2%) |  |
| This DOES NOT exist | 14 (51.9%) | 37 (56.1%) | 51 (54.8%) |  |
| I am not sure | 7 (25.9%) | 11 (16.7%) | 18 (19.4%) |  |
| Not applicable at my workplace | 2 (7.4%) | 1 (1.5%) | 3 (3.2%) |  |
| **changed post-test counseling note documentation** |  |  |  |  |
| This exists and a GC was involved | 5 (18.5%) | 10 (15.2%) | 15 (16.1%) |  |
| This exists and a GC was NOT involved | 0 (0.0%) | 0 (0.0%) | 0 (0.0%) |  |
| This exists and I don’t know if a GC was involved | 0 (0.0%) | 2 (3.0%) | 2 (2.2%) |  |
| This DOES NOT exist | 13 (48.1%) | 37 (56.1%) | 50 (53.8%) |  |
| I am not sure | 8 (29.6%) | 15 (22.7%) | 23 (24.7%) |  |
| Not applicable at my workplace | 1 (3.7%) | 2 (3.0%) | 3 (3.2%) |  |
| **changed laboratory report language** |  |  |  |  |
| This exists and a GC was involved | 1 (3.7%) | 0 (0.0%) | 1 (1.1%) |  |
| This exists and a GC was NOT involved | 0 (0.0%) | 3 (4.5%) | 3 (3.2%) |  |
| This exists and I don’t know if a GC was involved | 0 (0.0%) | 0 (0.0%) | 0 (0.0%) |  |
| This DOES NOT exist | 12 (44.4%) | 33 (50.0%) | 45 (48.4%) |  |
| I am not sure | 12 (44.4%) | 23 (34.8%) | 35 (37.6%) |  |
| Not applicable at my workplace | 2 (7.4%) | 7 (10.6%) | 9 (9.7%) |  |
| **changed patient portal policies** |  |  |  | 0.156 |
| This exists and a GC was involved | 5 (18.5%) | 3 (4.5%) | 8 (8.6%) |  |
| This exists and a GC was NOT involved | 3 (11.1%) | 8 (12.1%) | 11 (11.8%) |  |
| This exists and I don’t know if a GC was involved | 2 (7.4%) | 12 (18.2%) | 14 (15.1%) |  |
| This DOES NOT exist | 6 (22.2%) | 14 (21.2%) | 20 (21.5%) |  |
| I am not sure | 11 (40.7%) | 24 (36.4%) | 35 (37.6%) |  |
| Not applicable at my workplace | 0 (0.0%) | 5 (7.6%) | 5 (5.4%) |  |

Supplementary Table 2. Perceived workflow impact, harms, and benefits, comparing ordering and non-ordering providers

| Due to the implementation of the Final Rule, I... | Ordering provider (N=24) | Non-ordering provider (N=55) | Total (N=79) | p value (trend test for ordinal variables) |
| --- | --- | --- | --- | --- |
| ***Workflow impact*** |  |  |  |  |
| **need to check the electronic medical record and/or my email more frequently** |  |  |  | 0.984 |
| Strongly disagree | 2 (8.3%) | 4 (7.3%) | 6 (7.6%) |  |
| Disagree | 4 (16.7%) | 11 (20.0%) | 15 (19.0%) |  |
| Neither agree nor disagree | 4 (16.7%) | 10 (18.2%) | 14 (17.7%) |  |
| Agree | 10 (41.7%) | 19 (34.5%) | 29 (36.7%) |  |
| Strongly agree | 4 (16.7%) | 11 (20.0%) | 15 (19.0%) |  |
| **have less time to review results before discussing with a patient** |  |  |  | 0.263 |
| Strongly disagree | 2 (8.3%) | 3 (5.5%) | 5 (6.3%) |  |
| Disagree | 4 (16.7%) | 4 (7.3%) | 8 (10.1%) |  |
| Neither agree nor disagree | 1 (4.2%) | 3 (5.5%) | 4 (5.1%) |  |
| Agree | 12 (50.0%) | 21 (38.2%) | 33 (41.8%) |  |
| Strongly agree | 5 (20.8%) | 24 (43.6%) | 29 (36.7%) |  |
| **receive increased patient communications prior to my results review and disclosure** |  |  |  | 0.530 |
| Strongly disagree | 0 (0.0%) | 4 (7.3%) | 4 (5.1%) |  |
| Disagree | 4 (16.7%) | 12 (21.8%) | 16 (20.3%) |  |
| Neither agree nor disagree | 4 (16.7%) | 8 (14.5%) | 12 (15.2%) |  |
| Agree | 12 (50.0%) | 18 (32.7%) | 30 (38.0%) |  |
| Strongly agree | 4 (16.7%) | 13 (23.6%) | 17 (21.5%) |  |
| **changed my workflow to comply with the Final Rule** |  |  |  | 0.768 |
| Strongly disagree | 0 (0.0%) | 4 (7.3%) | 4 (5.1%) |  |
| Disagree | 7 (29.2%) | 13 (23.6%) | 20 (25.3%) |  |
| Neither agree nor disagree | 5 (20.8%) | 14 (25.5%) | 19 (24.1%) |  |
| Agree | 9 (37.5%) | 17 (30.9%) | 26 (32.9%) |  |
| Strongly agree | 3 (12.5%) | 7 (12.7%) | 10 (12.7%) |  |
| **face coordination of care challenges within my department example results going to a geneticist first delaying my review of results** |  |  |  | 0.931 |
| Strongly disagree | 4 (16.7%) | 8 (14.5%) | 12 (15.2%) |  |
| Disagree | 11 (45.8%) | 20 (36.4%) | 31 (39.2%) |  |
| Neither agree nor disagree | 4 (16.7%) | 11 (20.0%) | 15 (19.0%) |  |
| Agree | 3 (12.5%) | 10 (18.2%) | 13 (16.5%) |  |
| Strongly agree | 2 (8.3%) | 6 (10.9%) | 8 (10.1%) |  |
| **face coordination of care challenges with providers outside of my department example patients seeing results and messaging non-ordering providers for interpretation before I can discuss** |  |  |  | 0.220 |
| Strongly disagree | 3 (12.5%) | 4 (7.3%) | 7 (8.9%) |  |
| Disagree | 3 (12.5%) | 17 (30.9%) | 20 (25.3%) |  |
| Neither agree nor disagree | 2 (8.3%) | 8 (14.5%) | 10 (12.7%) |  |
| Agree | 13 (54.2%) | 17 (30.9%) | 30 (38.0%) |  |
| Strongly agree | 3 (12.5%) | 9 (16.4%) | 12 (15.2%) |  |
| **have one less administrative task because results are sent to patients automatically** |  |  |  | 0.470 |
| Strongly disagree | 5 (20.8%) | 12 (21.8%) | 17 (21.5%) |  |
| Disagree | 13 (54.2%) | 22 (40.0%) | 35 (44.3%) |  |
| Neither agree nor disagree | 1 (4.2%) | 8 (14.5%) | 9 (11.4%) |  |
| Agree | 3 (12.5%) | 11 (20.0%) | 14 (17.7%) |  |
| Strongly agree | 2 (8.3%) | 2 (3.6%) | 4 (5.1%) |  |
| ***Harms*** | **Ordering provider (N=27)** | **Non-ordering provider (N=63)** | **Total (N=90)** | **p value** |
| **could have strong emotional reactions when reviewing results notes on their own** |  |  |  | 0.002 |
| Disagree | 3 (11.1%) | 0 (0.0%) | 3 (3.3%) |  |
| Neither agree nor disagree | 3 (11.1%) | 0 (0.0%) | 3 (3.3%) |  |
| Agree | 12 (44.4%) | 31 (49.2%) | 43 (47.8%) |  |
| Strongly agree | 9 (33.3%) | 32 (50.8%) | 41 (45.6%) |  |
| **could misunderstand or misinterpret results** |  |  |  | 0.025 |
| Disagree | 1 (3.7%) | 0 (0.0%) | 1 (1.1%) |  |
| Neither agree nor disagree | 0 (0.0%) | 1 (1.6%) | 1 (1.1%) |  |
| Agree | 14 (51.9%) | 18 (28.6%) | 32 (35.6%) |  |
| Strongly agree | 12 (44.4%) | 44 (69.8%) | 56 (62.2%) |  |
| **could contact the inappropriate party to discuss results (example: direct laboratory or non-ordering provider contact)** |  |  |  | 0.023 |
| Strongly disagree | 1 (3.7%) | 0 (0.0%) | 1 (1.1%) |  |
| Disagree | 2 (7.4%) | 1 (1.6%) | 3 (3.3%) |  |
| Neither agree nor disagree | 3 (11.1%) | 6 (9.5%) | 9 (10.0%) |  |
| Agree | 16 (59.3%) | 26 (41.3%) | 42 (46.7%) |  |
| Strongly agree | 5 (18.5%) | 30 (47.6%) | 35 (38.9%) |  |
| ***Benefits*** | **Ordering provider (N=27)** | **Non-ordering provider (N=64)** | **Total (N=91)** | **p value** |
| **have more knowledge/context/questions at our disclosure session due to previous results review** |  |  |  | 0.007 |
| Strongly disagree | 0 (0.0%) | 1 (1.6%) | 1 (1.1%) |  |
| Disagree | 4 (15.4%) | 7 (10.9%) | 11 (12.2%) |  |
| Neither agree nor disagree | 0 (0.0%) | 15 (23.4%) | 15 (16.7%) |  |
| Agree | 20 (76.9%) | 29 (45.3%) | 49 (54.4%) |  |
| Strongly agree | 2 (7.7%) | 12 (18.8%) | 14 (15.6%) |  |
| **are empowered due to direct access to their medical records** |  |  |  | 0.233 |
| Strongly disagree | 0 (0.0%) | 1 (1.6%) | 1 (1.1%) |  |
| Disagree | 1 (3.7%) | 3 (4.7%) | 4 (4.4%) |  |
| Neither agree nor disagree | 3 (11.1%) | 20 (31.2%) | 23 (25.3%) |  |
| Agree | 18 (66.7%) | 32 (50.0%) | 50 (54.9%) |  |
| Strongly agree | 5 (18.5%) | 8 (12.5%) | 13 (14.3%) |  |
| **have increased engagement due to time to emotionally process results on their own before our conversation** |  |  |  | 0.057 |
| Strongly disagree | 1 (3.7%) | 0 (0.0%) | 1 (1.1%) |  |
| Disagree | 4 (14.8%) | 14 (21.9%) | 18 (19.8%) |  |
| Neither agree nor disagree | 6 (22.2%) | 24 (37.5%) | 30 (33.0%) |  |
| Agree | 14 (51.9%) | 16 (25.0%) | 30 (33.0%) |  |
| Strongly agree | 2 (7.4%) | 10 (15.6%) | 12 (13.2%) |  |
| **could have earlier reassurance about results** |  |  |  | 0.185 |
| Disagree | 2 (7.4%) | 2 (3.1%) | 4 (4.4%) |  |
| Neither agree nor disagree | 2 (7.4%) | 16 (25.0%) | 18 (19.8%) |  |
| Agree | 19 (70.4%) | 36 (56.2%) | 55 (60.4%) |  |
| Strongly agree | 4 (14.8%) | 10 (15.6%) | 14 (15.4%) |  |
| **could take earlier action on results** |  |  |  | 0.715 |
| Strongly disagree | 2 (7.4%) | 4 (6.2%) | 6 (6.6%) |  |
| Disagree | 4 (14.8%) | 14 (21.9%) | 18 (19.8%) |  |
| Neither agree nor disagree | 9 (33.3%) | 23 (35.9%) | 32 (35.2%) |  |
| Agree | 10 (37.0%) | 15 (23.4%) | 25 (27.5%) |  |
| Strongly agree | 2 (7.4%) | 8 (12.5%) | 10 (11.0%) |  |
| **have easier access to share their results chart with other providers without genetic counselor involvement** |  |  |  | 0.305 |
| Strongly disagree | 2 (7.4%) | 1 (1.6%) | 3 (3.3%) |  |
| Disagree | 1 (3.7%) | 2 (3.1%) | 3 (3.3%) |  |
| Neither agree nor disagree | 1 (3.7%) | 10 (15.6%) | 11 (12.1%) |  |
| Agree | 13 (48.1%) | 31 (48.4%) | 44 (48.4%) |  |
| Strongly agree | 10 (37.0%) | 20 (31.2%) | 30 (33.0%) |  |

Supplemental Table 3. Qualitative code definitions and example quotes

| Domain | Codes/ subcodes (N) | Example(s) |
| --- | --- | --- |
| 1. Negative impact (N=41) |  |  |
| Patient emotional impact |  |  |
|  | Distress (9) | Patients calling or sending messages in distress the moment they see the results are available, when we have not had an opportunity to review ourselves/with the geneticist.    Parents of my patients often have increased anxiety about results. One patient had li fraumeni syndrome and parents found out the results on friday at 6pm and were unable to talk to someone until tuesday because it was a holiday weekend it caused them extreme emotional distress.    I have had multiple patients receive confusing, concerning, and frightening results through their portals. Often this is at a time of day or weekend when there is not a provider available to explain the results. I have had patients come in on Monday because of a result they got Friday night, having not slept and having spent the weekend trying to make heads or tails of reports. |
|  | Distress SUBCODE: Panic/terrified/ freaking out (7) | An exome result was released after work on Friday revealing a likely pathogenic variant in 2 AR genes and a pathogenic variant in a third AR gene. The family was terrified that their children had all three rare genetic conditions and were in tears when I reached them on Monday and said they hadn't slept all weekend after seeing their child had a life limiting condition (which they did NOT have).    We send our Huntington's testing to a very large reference laboratory. Patients have direct access to their results through that laboratory portal. Recently had a positive result released directly to the patient who called me in a panic because she did not understand the result...    For many of our patients, the Final Rule does not impact them in a meaningful way. But for a subset of patients, the outcome of the Final Rule is psychological torture. |
|  | Anxiety (4) | I have had multiple patients receive confusing, concerning, and frightening results through their portals. Often this is at a time of day or weekend when there is not a provider available to explain the results. I have had patients come in on Monday because of a result they got Friday night, having not slept and having spent the weekend trying to make heads or tails of reports.    individuals review/Google their own reports and are not able to have helpful conversations with their MD because they can't move on from their own internet searches. |
|  | Surprised/shocked (2) | My patients don't remember a lot of what was discussed in pre-test counseling after they are shocked by the result in the portal. |
|  | Anger/offense (2) | Had an instance where a predictive testing patient for a neurodegenerative condition got the results before I could schedule a results disclosure. During pretest counseling, I acknowledged that they may have the ability to see results before disclosure, but that I strongly discouraged viewing them. Despite this guidance, they were very angry during our follow up appointment (ex. why I didn't immediately call them with results, their confusion about what the results meant). It was an incredibly frustrating situation to navigate. |
|  | Grief/regret (2) | I've had people find out fetal sex following a loss or termination that didn't ever want that information. I feel this is VERY different from those keeping sex a secret for a reveal party or until birth, that last group would have found out eventually. Learning the sex can complicate grieving and lead to unnecessary emotional distress for those that didn't want to fetal sex following a loss or termination.  Many patients have told me they regretted learning of their major medical diagnosis on MyChart |
| Misunderstanding results |  |  |
|  | Normal/ carrier/ VUS read as positive (12) | I have patients who have carrier screening results return and they think that they themselves have that particular condition.    I have had patients who misinterpreted the reports who were leaning toward termination before I explained what the report actually meant and what was actually going on. For many of our patients, the Final Rule does not impact them in a meaningful way. But for a subset of patients, the outcome of the Final Rule is psychological torture    An exome result was released after work on Friday revealing a likely pathogenic variant in 2 AR genes and a pathogenic variant in a third AR gene. The family was terrified that their children had all three rare genetic conditions and were in tears when I reached them on Monday and said they hadn't slept all weekend after seeing their child had a life limiting condition (which they did NOT have). They had contacted their on-call physician who verified the child had all three conditions based on the report when really they were at most a carrier of the conditions.    Patients get high risk screening or abnormal screening results for their pregnancy prior to review by a provider or a GC. |
|  | “Dr. Google” (4) | Patient received CDH1 VUS result over a weekend. spent the entire weekend researching and googling.    I work with patients in rural communities who only have a high school education and very low health literacy. However, they have access to go google. |
|  | Positive read as normal (3) | I had one case where a patient result was released but they had only viewed 1 page (just BRCA) which was negative. They missed the full panel which was a few pages in, which had a pathogenic variant in another gene. They were confused and thought the results were discordant |
|  | Poorly formatted reports (2) | I have had poorly formatted and difficult to read microarray results released to a patient before they were sent to me. |
|  | Other (4) - includes general references to misunderstanding and specific examples not easily categorized | Patients get results which are unclear or which they cannot interpret. |
| Contacting inappropriate provider |  |  |
|  | Contact lab instead of ordering physician (1) | I work in the lab and we get more patients calling the lab directly wanting to discuss results or testing family members because they see lab's phone number on the lab results before they have a chance to talk with their healthcare provider |
|  | Contact non-genetics provider (3) | I have had patients send their results to other healthcare providers without any explanation of what the result means sometimes including the patient's own interpretation of the result. |
| Patient harms or inconveniences |  |  |
|  | Receiving results in weekend/night (8) | One patient had li fraumeni syndrome and parents found out the results on friday at 6pm and were unable to talk to someone until tuesday because it was a holiday weekend it caused them extreme emotional distress    Patient received CDH1 VUS result over a weekend. spent the entire weekend researching and googling. Monday morning call with patient was spent mainly reassuring about VUS and the info patient had been reading    I have a prenatal patient who had a fetal MRI on a Friday morning. The report was signed and released Friday end of day and that's how she found out her baby has lissencephaly. We had already set up an interdisciplinary visit to review the results prior to their release for the next week. How devastating is that? She had to sit with that information for an entire weekend with no one to contact. |
|  | Serious news delivered by portal (6) | I have seen the most negative impact from automatic release of cfDNA results. Many patients have expressed significant distress about finding out about a positive result from an online portal rather than a phone call with a provider.    People find out they have cancer over an application. Access to care after reading these results is difficult and creates emotional distress |
|  | Danger of inappropriate care (2) | Another family found out about a VUS in the WT1 gene. They then transfered care to a different hospital and told them that the patient had deny's drash and patient almost underwent a radical nephrectomy before I was able to talk with parents/their new provider    I have had patients who misinterpreted the reports who were leaning toward termination before I explained what the report actually meant and what was actually going on. |
|  | Pt confused by clinic note (1) | While it hasn't happened to me personally, other providers have had patients complain that they don't understand what our notes mean (not recognizing that these notes are primarily meant to communicate with other healthcare providers). |
|  | Pt offended by clinic note (1) | We have had patients take offense to medical shorthand like SOB for shortness of breath. Or MTHFR for a gene name. |
|  | Pt learning unwanted info (1) | I've had people find out fetal sex following a loss or termination that didn't ever want that information. I feel this is VERY different from those keeping sex a secret for a reveal party or until birth, that last group would have found out eventually. Learning the sex can complicate grieving and lead to unnecessary emotional distress for those that didn't want to fetal sex following a loss or termination. |
| Workflow/ provider challenges |  |  |
|  | Not enough time for providers to review results (6) | Patients calling or sending messages in distress the moment they see the results are available, when we have not had an opportunity to review ourselves/with the geneticist. |
|  | Workflow disruptions (5) | One of my patients found out she was positive for both a BRCA1 pathogenic variant and a BRCA2 pathogenic variant, while I was in the middle of clinic. I fortunately had a no show during that time, but given the complex results, I felt the need to call the patient immediately. It is tough with conversations about complex results that may take up to an hour and how that can impact the rest of my day's agenda when patients randomly find out results. |
| No negative examples | No negative examples (3) | None. While some patients may read their results immediately, they also know to reach out to me to discuss them, so they don't have a lot of time to stress about results. |
| 2. Positive Impact (N=34) |  |  |
| Patient impact |  |  |
|  | Patients are more prepared for GC visit (8) | It does allow processing time for those that understand their results and receive them in advance. For those patients it can make our initial interaction following the result more productive. |
|  | Reassurance (7) | automatic release benefits my most anxious patients. due to work hours and clinic schedules, I cannot always immediately call out results. patients whose anxiety is relieved by simply having a result (good or bad news) can receive a result as soon as it is available rather than as soon as I am available.    Immediate release of negative test results has decreased stress and anxiety of waiting for a phone call or provider message to review results. |
|  | Empowers patients (2) | It empowers my patients to act as partners in their own medical care - they know that I am watching results, but that they can watch for results too. It allows families to better chose where and when they want to receive results, outside of the typical medical model of results disclosure appointments. |
|  | Easier for patients to access/share results and access care (5) | I work in adult oncology where results can impact treatment and it is helpful for oncologists to be able to see the results in case they are meeting with the patient before I can talk the the patient about their results.    The only positive impact has been patients ability to show me outside provider notes and records if I don't have them. However, this could easily be done without the Final Rule    the patient doesn't need to wait for their ordering provider to refer them for follow up GC to review the results now; they can call me directly to set up GC when their results are out. Patients get follow up GC faster because of this rule. |
|  | Patients get results sooner (6) | Patients receive normal results about their pregnancy in a quicker time frame |
|  | Sharing information with family (4) | Patients are able to see their results immediately, so they don't have to wait for results via physical mail. This is very helpful when it comes to efficiently scheduling screenings and testing for family members. |
|  | Reducing “lost” results (3) | Genetic tests ordered without genetic counselor involvement were released to a patient 2 years after they resulted. The patient was then able to reach out to a genetic counselor for help interpreting results and making medical decisions    if a provider forgets to call out a result, the patient will know still. |
|  | No positive examples (2) |  |
| Provider impact |  |  |
|  | Helps GCs work efficiently (11) | We have fewer patients "bugging" us for results because they can see themselves that the result is just not ready yet, and that it's not that we are "sitting on it."    One less thing to do, so I like the automatic results release.    if results are negative and I can see that the patient has accessed them, I feel that I do not have to respond as urgently as I might otherwise have. |
| Demographic characteristics (N=31) |  |  |
| Internet, tech, portal access |  |  |
|  | Low tech literacy (2) | Many are older adults who may not be tech savvy. |
|  | High tech literacy (1) | I work with a largely well educated population that is very tech savvy. But even this population has struggles with this rule. |
|  | Limited tech/portal access (9) | Not all patients have MyChart or ready access to it (d/t not having a web-enabled device or easy access to it). While most patients do have a web-enabled phone, we are failing our patients without this, and these patients are typically lower SES and receive poorer care already.    Most of my patients choose not to use the EHR because they either have low computer literacy or do not have reliable internet access. So the new rules only impact a subset of my patient population. Honestly the people who don't use our EHR would probably be more likely to experience the negative side of the rules (fear,misinterpretation,etc). |
| Health literacy and education |  |  |
|  | Low edu/ health literacy (6) | Final Rule may not really help patient populations with low health literacy or poor electronic tool access / skills. They may not be able to see results electronically or they may not understand what the clinical meaning of lab result they are seeing without interpretation from their healthcare provider    My patient population has generally lower health literacy than national averages and I tend to see two distinct reactions from families. They either open results, don't understand what they are reading, and PANIC - or - they see that results are available and purposefully do not open them until they've spoken with their providers. |
|  | High edu/health literacy (3) | Most are highly educated, anxious and depressed. |
| SES/ underserved | General SES/underserved (6) | My patients are primarily lower socio economic population with lower health literacy. Many are POC who already have distrust in the system.    Our genetic counseling group serves individuals from a wide spectrum of socioeconomic status. There are certainly patients who have limited access to technology and may be unable to have near-immediate access to results released through the portal. |
|  | Limited English proficiency (7) | The medical record is not translated into other languages so this rule is limited for non english speakers or those that can read in english. |
| Other patient characteristics |  |  |
|  | Mental health or psychosocial concerns (3) | Usually the most anxious parents are the ones that you prefer would not learn of their results from "My chart" versus from the gc. |
|  | Age (2) | I find younger individuals tend to ask for immediate access or utilization of portals over older populations. |
|  | Disabilities (1) | Many have significant visual impairments |
|  | Personal interest (1) | I think people with a lot of interest in genetics (ex. physicians, nurses, other STEM folk) may be inclined to review it on their own earlier because they have perhaps a better understanding or higher health literacy to read the report and understand it. Other folks without that personal interest in genetics or with lower health literacy, may choose to wait to read until the result has been disclosed to them or read it and have more questions. |
| Additional codes |  |  |
|  | Disparities (5) – concern about widening health disparities | Not all patients have MyChart or ready access to it (d/t not having a web-enabled device or easy access to it). While most patients do have a web-enabled phone, we are failing our patients without this, and these patients are typically lower SES and receive poorer care already. |
|  | Clinical setting (5) | I work mostly with adult cardiology and would have much more significant concerns in some other practice settings.    as a cancer counselor, my patient population has far more complaints about learning of their cancer diagnosis from automatic results than they do about the genetic results being released automatically.    I work in prenatal where everything is time sensitive and often highly emotional. I've seen people from all different backgrounds negatively impacted by the final rule.    I am in a small, private company and results are not immediately available to the patient on our end as we do not have a patient portal. We do send results to the physician once available which I have heard from patients that they have gotten results that way before the consultation    As mentioned, immediate release of results is not at all appropriate for the adult neurological predictive testing populations where there are careful protocols in place for a reason. Final rule throws some of that out of the window. |
|  | Distrust (1) | My patients are primarily lower socio economic population with lower health literacy. Many are POC who already have distrust in the system. |
|  | “Regardless” (2) – Final Rule poses challenges even for non-disadvantaged patients | I work in prenatal where everything is time sensitive and often highly emotional. I've seen people from all different backgrounds negatively impacted by the final rule.    I work with a largely well educated population that is very tech savvy. But even this population has struggles with this rule. |
|  | None (1) | I have not noticed any population influences. |
| Practice changes (N=34) |  |  |
|  | have less time to review results before discussing with a patient. (4) | I do find that I need to call patients sooner and sometimes with less information and my conversations now sometimes include something to the effect of "I have your results but I need to do more research and will call you when I have more data."    I try as hard as possible to call out even negative results the same day they are released. |
|  | have one less administrative task because results are sent to patients automatically. (2) | It is nice though, for negative results, to not have to make multiple phone attempts when I can tell they saw the report. |
| Other codes |  |  |
|  | Pre-test counseling: SUBCODE Inform pt that they may get results before the GC or need time to interpret (18) | I now have to explicitly explain to patients that there are two timeframes: the turnaround time of the test and the time it takes to interpret the results in clinical context. I also have to remind patients that I will call them after both of these things have taken place so they may see their result a few weeks before hearing from me.    Consenting process has changed. I now give a heads-up to all patients that they may get results at the same time or even before me, on the weekends. |
|  | Pre-test counseling: SUBCODE Counsel to consider waiting to open results (6) | I now tell patients ahead of time in my pretest counseling that results will automatically be released to them so that they can brace themselves and decide if they want to look at the results on their own or wait for my phone call.    Encourage patients to wait to view results until after I am able to call them.    I let them know they can choose to open or not but we would discuss them thoroughly at the appointment. |
|  | Pre-test counseling: SUBCODE Consent to block automatic release (1) | I now talk with patients about if they want results automatically released or not, especially if I am particularly suspicious results will be positive (ie if ultrasound is highly suggestive of T18 in someone who is advanced age). |
|  | Pre-test counseling: SUBCODE  How results will arrive (4) | We discuss how disclosure happens within a certain timeframe of receiving the result, set expectations for the disclosure ahead of time to mitigate emotional stress.    I always make sure to explain the result delivery process as part of the pre-test counseling, which I think really avoids negative impacts |
|  | Documentation: SUBCODE Considerations for sensitive pt info (3) | Mainly we are careful what goes into a GC note. It was top of mind even before the Final Rule so I would not say it changed our workflow with writing notes etc, BUT just keeping it in mind always that the note you write actually reflects a person.    Almost af our communication internally is also visible in the chart. For example if I route a telephone call and add comments for the recipient, the patient can see. I've forgotten that and had to go back and delete it. It's difficult to comment on the psychosocial state of a patient, so many times I leave it out, which is a disservice to other providers. This is especially true when there is concerning behavior. Patients have asked me to leave out details like termination of pregnancies and even family history details. |
|  | Documentation: Other changes (4) | Prior to the final rules we had already implemented EPIC templates to improve efficiency of note writing so that consult notes and results letters can be released same day. We decided to continue to write a separate patient friendly results letter even though the patients can see everything their referring doctor gets. We have concerns about about this our patient population's medical/genetic literacy.    Document and discuss results being released to patient at same time they are received by clinical team and needing time for interpretation    As part of pre-test counseling we now standardly tell them, and have in the note that the results may be available in their record before we have had a chance to review, and we may not be immediately available to review results with them    We were given a tip sheet on documentation to be more patient friendly. I think it's good for us to think about how we write things and make language accessible. But you're asking doctors to see 40 pts in a day and also think critically about their note? That's burn out. |
|  | EHR portal (3) | We have to manually put in these results into the chart so the only way to truly grant immediate access is through the lab's patient portal. We offer this option up front instead of waiting for results to come in. We discuss how we manually put in these results so immediate access truly isn't feasible due to this step through our EHR and portal.    We are able to withhold ONLY Huntington disease test results, all other results are automatically released if the testing lab interfaces with our EMR. |
|  | Lab portal (2) | but working with labs like Invitae is difficult because they release to a patient before i even know the results are back      We have to manually put in these results into the chart so the only way to truly grant immediate access is through the lab's patient portal. We offer this option up front instead of waiting for results to come in. We discuss how we manually put in these results so immediate access truly isn't feasible due to this step through our EHR and portal. |
|  | Changes to lab GC role (1) | I also call more prelim results to my clinical GC colleagues so that they have a chance to call patients with abnormal results before they are release to the portal. |
|  | Changes to lab report (2) | I work in the lab and do report writing. Our lab decided to add a disclaimer at the end of the report for families that let them know they may be seeing the result before their healthcare providers, and if they have a question to contact the healthcare provider (and not the lab) to discuss results    I'm a laboratory GC - the draft results reports I write are different now, because I know patients will see them in their entirety and possibly before the ordering provider. |
| No changes | No changes (1) | I've been practicing since 2018 all at the same institution and I think patients have had access to their chart notes that entire time |
| Cross-cutting themes (N=4) |  |  |
| Potential solutions/ recommendations | Potential solutions/ recommendations | They need someone to call who gives immediate feedback when they have questions. Just enough info to relieve anxiety before their next visit with the ordering provider.    I am all for release of results to patients. I just wish there was an option for a three day delay to give providers a chance to contact the patient first.    I think there are some patient demographics who are more likely to use the benefits of the Final Rule than others so we should just be mindful of that in office policies regarding release of results to make sure those less likely to use the benefits still get their results.    immediate release of results is not at all appropriate for the adult neurological predictive testing populations where there are careful protocols in place for a reason. Final rule throws some of that out of the window. I think it can be harmful to highly anxious patients or those with low levels of health literacy. |
| Non-genetics results |  |  |
|  | Non-genetics results impact GC practice (1) | Numerous patients have told me how anxious and confused they are by trying to interpret various imaging, path and lab reports received before having a medical provider explains it to them    patients often complain about their inability to understand results that are automatically released examples include mammogram reports, pathology reports, MRI reports, etc. |
|  | Anti-exceptionalism (2) | as a cancer counselor, my patient population has far more complaints about learning of their cancer diagnosis from automatic results than they do about the genetic results being released automatically.    Patients receive all other results immediately - including other sensitive results such as dx of cancer and pregnancies related tests. So having genetic tests be treated like other lab tests is a step in the right direction, in my opinion, as we continue to see genetics move into the mainstream of cre. |
