## Supplementary material for "Navigating the Impact of the 21st Century Cures Act Final Rule: A National Cross-Sectional Survey of U.S. Genetic Counselors": survey code book

### Data Dictionary Codebook

02/27/2024 2:19pm

| # | Variable / Field Name | Field Label<br>*Field Note* | Field Attributes (Field Type, Validation, Choices, Calculations, etc.) |
|---|---|---|---|
| **Instrument: Consent Page (consent_page)** Enabled as survey | | | |
| 1 | [record_id1] | Record ID 1 | text |
| 2 | [inclusion] | During any point after April 2021, were you a practicing genetic counselor who worked in the United States? | yesno, Required<br>1 Yes<br>0 No<br>Stop actions on 0 |
| 3 | [consent_answer] | Do you consent to participate in this study? | yesno, Required<br>1 Yes<br>0 No<br>Stop actions on 0 |
| 4 | [consent_page_complete] | Section Header: *Form Status*<br>Complete? | dropdown<br>0 Incomplete<br>1 Unverified<br>2 Complete |
| **Instrument: Survey (survey)** Enabled as survey | | | |
| 5 | [finalruledefinition] | This survey is referencing the Final Rule of the 21st Century Cures Act (Final Rule). The Final Rule went into effect in April 2021 and required results and notes in the electronic medical record be available to patients without delay. There are some exceptions to this requirement. Implementation of the Final Rule may be institution specific. | descriptive |
| 6 | [training] | Did you complete training about the Final Rule prior to or shortly after implementation of the rule? (select all that apply) | checkbox<br>1 training___1 Training completed through my workplace before implementation<br>2 training___2 Training completed through my workplace after implementation<br>3 training___3 Training completed through continuing education opportunities outside of my workplace<br>4 training___4 Training completed through my genetic counseling program<br>5 training___5 I did not complete training<br>6 training___6 I am not sure |
| 7 | [contact] | Do you have a direct contact at your institution if you have questions about the Final Rule? | radio<br>1 Yes<br>2 No |

| | | | |
|---|---|---|---|
| | | | 3 Unsure |
| 8 | [state] | Do you know of a state law(s) in your state(s) of practice that have impacted the Final Rule implementation? | yesno<br>1 Yes<br>0 No |
| 9 | [state_details]<br>Show the field ONLY if:<br>[state] = '1' | Please provide details (if known) about the state laws. | notes |
| 10 | [policies_instituitional] | Section Header: *For each of the following topics, please indicate if it is present at your current workplace and if a genetic counselor (GC) at your workplace was involved in development/implementation of the resource. Mark the most appropriate option. Due to the Final Rule, my workplace has...*<br>institutional level policies for results/notes release. | radio (Matrix)<br>1 This exists and a GC was involved<br>2 This exists and a GC was NOT involved<br>3 This exists and I don't know if a GC was involved<br>4 This DOES NOT exist<br>5 I am not sure<br>6 Not applicable at my workplace |
| 11 | [policies_dept] | department, division, or workgroup level policies for results/notes release. | radio (Matrix)<br>1 This exists and a GC was involved<br>2 This exists and a GC was NOT involved<br>3 This exists and I don't know if a GC was involved<br>4 This DOES NOT exist<br>5 I am not sure<br>6 Not applicable at my workplace |
| 12 | [workgroup] | an established work group for Final Rule implementation and optimization. | radio (Matrix)<br>1 This exists and a GC was involved<br>2 This exists and a GC was NOT involved<br>3 This exists and I don't know if a GC was involved<br>4 This DOES NOT exist<br>5 I am not sure<br>6 Not applicable at my workplace |
| 13 | [pointperson] | a point person for questions about Final Rule/release of results/notes (provider or patient facing). | radio (Matrix)<br>1 This exists and a GC was involved<br>2 This exists and a GC was NOT involved<br>3 This exists and I don't know if a GC was involved<br>4 This DOES NOT exist<br>5 I am not sure<br>6 Not applicable at my workplace |
| 14 | [website] | a website on the Final Rule/release of results/notes (provider or patient facing). | radio (Matrix)<br>1 This exists and a GC was involved<br>2 This exists and a GC was NOT involved<br>3 This exists and I don't know if a GC was involved<br>4 This DOES NOT exist |

| | | | |
|---|---|---|---|
| | | | 5 I am not sure<br>6 Not applicable at my workplace |
| 15 | [tipsheet] | tipsheet(s) on the Final Rule/release of results/notes (provider or patient facing). | radio (Matrix)<br>1 This exists and a GC was involved<br>2 This exists and a GC was NOT involved<br>3 This exists and I don't know if a GC was involved<br>4 This DOES NOT exist<br>5 I am not sure<br>6 Not applicable at my workplace |
| 16 | [blocking] | a system to block results/notes release when aligned with the Final Rule. | radio (Matrix)<br>1 This exists and a GC was involved<br>2 This exists and a GC was NOT involved<br>3 This exists and I don't know if a GC was involved<br>4 This DOES NOT exist<br>5 I am not sure<br>6 Not applicable at my workplace |
| 17 | [pre_test] | changed pre-test counseling note documentation. | radio (Matrix)<br>1 This exists and a GC was involved<br>2 This exists and a GC was NOT involved<br>3 This exists and I don't know if a GC was involved<br>4 This DOES NOT exist<br>5 I am not sure<br>6 Not applicable at my workplace |
| 18 | [post_test] | changed post-test counseling note documentation. | radio (Matrix)<br>1 This exists and a GC was involved<br>2 This exists and a GC was NOT involved<br>3 This exists and I don't know if a GC was involved<br>4 This DOES NOT exist<br>5 I am not sure<br>6 Not applicable at my workplace |
| 19 | [labreport] | changed laboratory report language. | radio (Matrix)<br>1 This exists and a GC was involved<br>2 This exists and a GC was NOT involved<br>3 This exists and I don't know if a GC was involved<br>4 This DOES NOT exist<br>5 I am not sure<br>6 Not applicable at my workplace |
| 20 | [policies_portal] | changed patient portal policies. | radio (Matrix)<br>1 This exists and a GC was involved |

| | | | |
|---|---|---|---|
| | | | 2 This exists and a GC was NOT involved<br>3 This exists and I don't know if a GC was involved<br>4 This DOES NOT exist<br>5 I am not sure<br>6 Not applicable at my workplace |
| 21 | [ checking ] | Section Header: *Please select your level of agreement with each of the following statements. Due to the implementation of the Final Rule, I ...*<br>need to check the electronic medical record and/or my email more frequently. | radio (Matrix)<br>1 Strongly disagree<br>2 Disagree<br>3 Neither agree nor disagree<br>4 Agree<br>5 Strongly agree<br>6 Not applicable for my position |
| 22 | [ timing ] | have less time to review results before discussing with a patient. | radio (Matrix)<br>1 Strongly disagree<br>2 Disagree<br>3 Neither agree nor disagree<br>4 Agree<br>5 Strongly agree<br>6 Not applicable for my position |
| 23 | [ communications ] | receive increased patient communications prior to my results review and disclosure. | radio (Matrix)<br>1 Strongly disagree<br>2 Disagree<br>3 Neither agree nor disagree<br>4 Agree<br>5 Strongly agree<br>6 Not applicable for my position |
| 24 | [ change ] | changed my workflow to comply with the Final Rule. | radio (Matrix)<br>1 Strongly disagree<br>2 Disagree<br>3 Neither agree nor disagree<br>4 Agree<br>5 Strongly agree<br>6 Not applicable for my position |
| 25 | [ coordination_dept ] | face coordination of care challenges within my department (example, results going to a geneticist first delaying my review of results). | radio (Matrix)<br>1 Strongly disagree<br>2 Disagree<br>3 Neither agree nor disagree<br>4 Agree<br>5 Strongly agree<br>6 Not applicable for my position |
| 26 | [ coordination_other ] | face coordination of care challenges with providers outside of my department (example, patients seeing | radio (Matrix)<br>1 Strongly disagree |

| | | | |
|---|---|---|---|
| | | results and messaging non-ordering providers for interpretation before I can discuss). | 2 Disagree<br>3 Neither agree nor disagree<br>4 Agree<br>5 Strongly agree<br>6 Not applicable for my position |
| 27 | [ sending_results ] | have one less administrative task because results are sent to patients automatically. | radio (Matrix)<br>1 Strongly disagree<br>2 Disagree<br>3 Neither agree nor disagree<br>4 Agree<br>5 Strongly agree<br>6 Not applicable for my position |
| 28 | [ pt_reaction ] | Section Header: *Please select your level of agreement for each of the following statements. Due to the implementation of the Final Rule, I believe patients...*<br>could have strong emotional reactions when reviewing results/notes on their own. | radio (Matrix)<br>1 Strongly disagree<br>2 Disagree<br>3 Neither agree nor disagree<br>4 Agree<br>5 Strongly agree<br>6 Not applicable for my position |
| 29 | [ pt_understand ] | could misunderstand or misinterpret results. | radio (Matrix)<br>1 Strongly disagree<br>2 Disagree<br>3 Neither agree nor disagree<br>4 Agree<br>5 Strongly agree<br>6 Not applicable for my position |
| 30 | [ pt_contact ] | could contact the inappropriate party to discuss results (example: direct laboratory or non-ordering provider contact). | radio (Matrix)<br>1 Strongly disagree<br>2 Disagree<br>3 Neither agree nor disagree<br>4 Agree<br>5 Strongly agree<br>6 Not applicable for my position |
| 31 | [ pt_knowledge ] | have more knowledge/context/questions at our disclosure session due to previous results review. | radio (Matrix)<br>1 Strongly disagree<br>2 Disagree<br>3 Neither agree nor disagree<br>4 Agree<br>5 Strongly agree<br>6 Not applicable for my position |
| 32 | [ pt_empower ] | are empowered due to direct access to their medical records. | radio (Matrix)<br>1 Strongly disagree<br>2 Disagree |

| | | | |
|---|---|---|---|
| | | | 3 Neither agree nor disagree<br>4 Agree<br>5 Strongly agree<br>6 Not applicable for my position |
| 33 | [pt_processing] | have increased engagement due to time to emotionally process results on their own before our conversation. | radio (Matrix)<br>1 Strongly disagree<br>2 Disagree<br>3 Neither agree nor disagree<br>4 Agree<br>5 Strongly agree<br>6 Not applicable for my position |
| 34 | [pt_assurance] | could have earlier reassurance about results. | radio (Matrix)<br>1 Strongly disagree<br>2 Disagree<br>3 Neither agree nor disagree<br>4 Agree<br>5 Strongly agree<br>6 Not applicable for my position |
| 35 | [pt_action] | could take earlier action on results. | radio (Matrix)<br>1 Strongly disagree<br>2 Disagree<br>3 Neither agree nor disagree<br>4 Agree<br>5 Strongly agree<br>6 Not applicable for my position |
| 36 | [pt_share] | have easier access to share their results/chart with other providers without genetic counselor involvement. | radio (Matrix)<br>1 Strongly disagree<br>2 Disagree<br>3 Neither agree nor disagree<br>4 Agree<br>5 Strongly agree<br>6 Not applicable for my position |
| 37 | [examples_positive] | Section Header:<br>Please describe any examples you wish to share of how the Final Rule has positively impacted your patients/clients. | notes |
| 38 | [examples_negative] | Please describe any examples you wish to share of how the Final Rule has negatively impacted your patients/clients. | notes |
| 39 | [patients_characteristics] | Are there any demographics or characteristics of the population you typically work with that influence your perspective of the Final Rule (potential impact of patients having near-immediate access to their notes and results)? Please elaborate. | notes |
| 40 | [changes] | If you practiced prior to the implementation of the Final Rule (April 2021), please describe any examples you | notes |

| | | | |
|---|---|---|---|
| | | wish to share about how the Final Rule has changed your personal practice as a genetic counselor including any changes you've made to consenting, documentation, patient results release, patient discussions, client conversations, report writing, etc. | |
| 41 | [demo_age] | Section Header: *Demographics*<br>What is your age in years? | dropdown<br>1 \| 20-24<br>2 \| 25-29<br>3 \| 30-34<br>4 \| 35-39<br>5 \| 40-44<br>6 \| 45-49<br>7 \| 50-54<br>8 \| 55-59<br>9 \| 60-64<br>10 \| 65-69<br>11 \| 70 or older |
| 42 | [demo_gender] | What is your gender identity? (select all that apply) | checkbox<br>1 \| demo_gender___1 \| Man<br>2 \| demo_gender___2 \| Non-binary<br>3 \| demo_gender___3 \| Transgender<br>4 \| demo_gender___4 \| Woman<br>5 \| demo_gender___5 \| Other<br>8 \| demo_gender___8 \| Unsure/questioning<br>7 \| demo_gender___7 \| Prefer not to Say |
| 43 | [demo_gender_other]<br>Show the field ONLY if:<br>[demo_gender(5)] = '1' | Other gender identity: | text |
| 44 | [demo_raceethnicity] | What is your race and/or ethnicity? (select all that apply) | checkbox<br>1 \| demo_raceethnicity___1 \| African American or Black<br>2 \| demo_raceethnicity___2 \| Native American/ Alaska Native/ First Nations/ Indigenous<br>3 \| demo_raceethnicity___3 \| Asian<br>4 \| demo_raceethnicity___4 \| Native Hawaiian/ Pacific Islander<br>5 \| demo_raceethnicity___5 \| White<br>6 \| demo_raceethnicity___6 \| Other<br>7 \| demo_raceethnicity___7 \| Non-Hispanic or -Latino<br>8 \| demo_raceethnicity___8 \| Hispanic or Latino |
| 45 | [demo_raceethnicity_other]<br>Show the field ONLY if: | Other race or ethnicity: | text |

| | | | |
|---|---|---|---|
| | [demo_raceethnicity(6)] = '1' | | |
| 46 | [demo_list] | What geographic region do you work? | dropdown<br>1 Region 1 (CT, MA, ME, NH, RI, VT, CN Maritime Provinces)<br>2 Region 2 (DC, DE, MD, NJ, NY, PA, VA, WV, PR, VI, Quebec)<br>3 Region 3 (AL, FL, GA, KY, LA, MS, NC, SC, TN)<br>4 Region 4 (AR, IA, IL, IN, KS, MI, MN, MO, ND, NE, OH, OK, SD, WI, Ontario)<br>5 Region 5 (AZ, CO, MT, NM, TX, UT, WY, Alberta, Manitoba, Sask.)<br>6 Region 6 (AK, CA, HI, ID, NV, OR, WA, British Columbia) |
| 47 | [position] | What type of position do you currently work as your primary role? | radio<br>1 Direct patient care<br>2 Non-direct patient care<br>3 Mixed position |
| 48 | [ordering_provider]<br>Show the field ONLY if:<br>[position] = '1' or [position] = '3' | Are you an ordering provider for testing sent on your patients? | radio<br>1 Yes. I place and sign orders as the provider of record<br>2 No. I place the order but am not listed as the ordering provider<br>3 No. I cannot place or sign orders<br>4 No. I do not order in my role |
| 49 | [resultsretrieval]<br>Show the field ONLY if:<br>[position] = '1' and [position] = '3' | When a patient's results report is complete, how do you receive results? | checkbox<br>1 resultsretrieval___1 I do not receive results reports in my role<br>2 resultsretrieval___2 My Individual Work Email<br>3 resultsretrieval___3 Group Email (checked by others)<br>4 resultsretrieval___4 Group Email (checked by me)<br>5 resultsretrieval___5 Electronic Medical Record Basket directly<br>6 resultsretrieval___6 Electronic Medical Record Basket sent by another provider<br>7 resultsretrieval___7 Fax<br>8 resultsretrieval___8 Mail<br>9 resultsretrieval___9 I do not receive them directly and must seek them out. |
| 50 | [workplacesetting] | In what setting(s) do you currently work? (select all that apply) | checkbox<br>1 workplacesetting___1 Government Organization or Ager |

| | | | |
|---|---|---|---|
| | | | 2 workplacesetting___2 Hospital/Medical Facility- Academic Medical Center<br>3 workplacesetting___3 Hospital/Medical Facility- Public or Community Based Hospital (including FQHC)<br>4 workplacesetting___4 Hospital/Medical Facility- Private<br>5 workplacesetting___5 Insurance Company/Benefit Management Compa<br>6 workplacesetting___6 Laboratory Commerc<br>7 workplacesetting___7 Not-For-Profit Organization (non-hospital)<br>8 workplacesetting___8 Private Practice<br>9 workplacesetting___9 Private Company- Biotechnology/Resea development, Digital Health/Software, Pharmaceutical<br>10 workplacesetting___10 Other |
| 51 | [workplacesetting_other]<br>Show the field ONLY if: [workplacesetting(10)] = '1' | Other workplace setting: | text |
| 52 | [demo_years] | How many years experience do you have as a practicing genetic counselor (round to the nearest year)? | dropdown<br>1 < 1 year<br>2 1- 4 years<br>3 5 - 9 years<br>4 10 - 14 years<br>5 15 - 19 years<br>6 20 - 24 years<br>7 25 - 29 years<br>8 30 - 34 years<br>9 35 - 40 years<br>10 40+ years |
| 53 | [demo_yeargraduate] | What year did you graduate from a genetic counseling program? | radio<br>1 Before 2021<br>2 2021<br>3 2022 or 2023 |
| 54 | [demo_specialty] | Please indicate the specialty area(s) you have practiced in since April 2021. (select all that apply) | checkbox<br>1 demo_specialty___1 Cancer Genetics- Adult<br>2 demo_specialty___2 Cancer Genetics- Pedia<br>3 demo_specialty___3 Cardiology |

| | | |
|---|---|---|
| 4 | demo_specialty___4 | Consumer Genomics/I genomics |
| 5 | demo_specialty___5 | General Adult Genetic |
| 6 | demo_specialty___6 | Genomic Medicine |
| 7 | demo_specialty___7 | Hematology |
| 8 | demo_specialty___8 | Laboratory Sciences (molecular/cytogeneti testing/variant science |
| 9 | demo_specialty___9 | Metabolic Disease |
| 10 | demo_specialty___10 | Neurogenetics |
| 11 | demo_specialty___11 | Ophthalmology |
| 12 | demo_specialty___12 | Pediatrics |
| 13 | demo_specialty___13 | Preconception/Reprod screening |
| 14 | demo_specialty___14 | Preimplantation genet ART/IVF, infertility |
| 15 | demo_specialty___15 | Prenatal |
| 16 | demo_specialty___16 | Public Health |
| 17 | demo_specialty___17 | Other Specialty |

| | | | |
|---|---|---|---|
| 55 | [demo_specialty_other]<br>Show the field ONLY if:<br>[demo_specialty(17)] = '1' | Other specialty: | text |
| 56 | [survey_complete] | Section Header: *Form Status*<br>Complete? | dropdown<br>0 Incomplete<br>1 Unverified<br>2 Complete |
